# McQ – An open-source multiplexed SARS-CoV-2 quantification platform

**DOI:** 10.1101/2020.12.02.20242628

**Authors:** Sibylle C Vonesch, Danila Bredikhin, Nikolay Dobrev, Laura Villacorta, Rozemarijn Kleinendorst, Elisabetta Cacace, Julia Flock, Max Frank, Ferris Jung, Julia Kornienko, Karin Mitosch, Mireia Osuna-López, Jürgen Zimmermann, Stephan Göttig, Axel Hamprecht, Hans-Georg Kräusslich, Michael Knop, Athanasios Typas, Lars M Steinmetz, Vladimir Benes, Kim Remans, Arnaud R Krebs

**Affiliations:** European Molecular Biology Laboratory (EMBL), Genome Biology Unit, 69117, Heidelberg, Germany; European Molecular Biology Laboratory (EMBL), Protein Expression and Purification Facility, 69117, Heidelberg, Germany; European Molecular Biology Laboratory (EMBL), Genomics Core Facility, 69117, Heidelberg, Germany; Institute of Medical Microbiology and Infection Control, Hospital of Goethe University, Frankfurt am Main, Germany; Institut für medizinische Mikrobiologie, Immunologie und Hygiene, 50935 Köln, Germany; Institut für med. Mikrobiologie und Virologie, Carl von Ossietzky Universität Oldenburg, 26129 Oldenburg, Germany; Center for Infectious Diseases, Department of Virology, Universitätsklinikum Heidelberg, 69117, Heidelberg, Germany; Deutsches Zentrum für Infektionsforschung, partner site Heidelberg, Heidelberg, Germany; Center for Molecular Biology of Heidelberg University (ZMBH), Heidelberg, Germany; German Center for Infection Research (DZIF), Heidelberg, Germany; German Cancer Research Center (DKFZ), Heidelberg, Germany; DKFZ-ZMBH Alliance, Heidelberg, Germany; Stanford Genome Technology Center, Stanford University, 3165 Porter Drive, Palo Alto, CA 94304, USA; Department of Genetics, Stanford University School of Medicine, Stanford, California, USA

## Abstract

McQ is a SARS-CoV-2 quantification assay that couples early-stage barcoding with high-throughput sequencing to enable multiplexed processing of thousands of samples. McQ is based on homemade enzymes to enable low-cost testing of large sample pools, circumventing supply chain shortages.

Implementation of cost-efficient high-throughput methods for detection of RNA viruses such as SARS-CoV-2 is a potent strategy to curb ongoing and future pandemics. Here we describe Multiplexed SARS-CoV-2 Quantification platform (McQ), an in-expensive scalable framework for SARS-CoV-2 quantification in saliva samples. McQ is based on the parallel sequencing of barcoded amplicons generated from SARS-CoV-2 genomic RNA. McQ uses indexed, target-specific reverse transcription (RT) to generate barcoded cDNA for amplifying viral- and human-specific regions. The barcoding system enables early sample pooling to reduce hands-on time and makes the ap-proach scalable to thousands of samples per sequencing run. Robust and accurate quantification of viral load is achieved by measuring the abundance of Unique Molecular Identifiers (UMIs) introduced during reverse transcription. The use of homemade reverse transcriptase and polymerase enzymes and non-proprietary buffers reduces RNA to library reagent costs to 92 cents/sample and circumvents potential supply chain short-ages. We demonstrate the ability of McQ to robustly quantify various levels of viral RNA in 838 clinical samples and accu-rately diagnose positive and negative control samples in a test-ing workflow entailing self-sampling and automated RNA ex-traction from saliva. The implementation of McQ is modular, scalable and could be extended to other pathogenic targets in future.

## Introduction

The coronavirus disease 2019 (COVID-19) has created a worldwide pandemic with over 59 million reported cases and1.4 million recorded deaths worldwide ^1^. While population-wide lockdown measures help to contain the rapid spread of the virus, thereby alleviating the burden on the health care system, they come at a severe economic and societal cost. Thus, lockdowns are not sustainable and alternative measures have been proposed to enable social and economic life to resume in the absence of an effective vaccine. These include social distancing measures and wearing of face protection in public spaces. Another important leverage in the fight to control the pandemic is increasing testing capacity to enable systematic surveillance testing, which combined with contact tracing and rapid isolation of infectious individuals can curtail viral spread ^2,3^.

One of the challenges in the control of the COVID-19 pandemic is the high frequency of pre- or asymptomatic individuals that nevertheless display high levels of infectivity, making containment of transmission solely by symptomatic testing impossible ^4–6^. This is the case for a majority of patients during the 2-7 days of the incubation phase ^7^ and lasts during the entire duration of the infection for approximately one fifth of individuals ^8^. Detection of these asymptomatic spreaders could be achieved by the implementation of high-frequency population-scale surveillance testing. This would imply testing all members of a community repeatedly and regardless of symptoms. Achieving such a goal requires the development of scalable testing strategies that are accessible and cost efficient.

The gold standard COVID-19 diagnosis is by quantitative RT-qPCR against the viral genomic RNA that is contained in nasal or throat swab samples. The assay is highly sensitive as it can detect very low abundances of viral particles. Current challenges include the use of commercial reagents that are subject to shortage, poor scalability and the high cost of the test, which preclude its adoption for population-scale testing of asymptomatic individuals. To circumvent these limitations, several alternative technologies have been developed ^9^. These are based on the detection of antigens ^10–13^, colorimetric assays ^14–18^, CRISPR-based detection ^19–22^ or multiplexed detection of nucleic acids using NGS^23–29^.

Here we describe the development of an integrated high-throughput testing workflow for multiplexed SARS-CoV-2 quantification (McQ). The strategy entails parallel sequencing of barcoded amplicons generated from SARS-CoV-2 genomic RNA. Samples are barcoded by targeted reverse transcription against viral and human target regions to label each sample uniquely and enable pooling for barcoded amplification. Short read Illumina sequencing is used to quantify viral abundance in individual samples, using unique molecular identifiers added during reverse transcription for improved robustness and reduced variability in viral load quantification. Quality control, data analysis and diagnostics are performed using public open source code. To circumvent reagent shortages McQ largely relies on homemade enzymes and non-commercial buffers. We benchmarked McQ against existing technologies, and validated its performance through the parallel testing of >800 clinical samples. We combined McQ with automated RNA extraction from saliva samples from healthy volunteers and control samples. We confirm McQ diagnostics of all saliva samples as negative with LAMP and RT-qPCR, and show accurate diagnostics of positive and negative controls.

## Results

A key feature to enable scalability of SARS-CoV-2 genomic RNA quantification resides in lowering the per sample hands-on time. To that end, we adapted a reverse transcription barcoding strategy ^25^ to enable early sample pooling and parallelised processing. This enables scaling to thousands of samples per sequencing run. McQ uses Unique Molecular Identifiers (UMIs) to correct for PCR amplification bias, enabling more accurate quantification of viral load. The McQ sample preparation workflow relies mostly on homemade enzymes and reagents to circumvent supply chain shortages and to lower testing cost.

### McQ experimental design

McQ is based on the parallel sequencing of barcoded amplicons generated from SARS-CoV-2 genomic RNA (Figure 1). McQ was established using RNA purified from clinical swab samples or saliva samples provided by healthy volunteers. We implemented an automated extraction workflow based on magnetic beads and non-proprietary buffers leading to good RNA yield and purity (Supplementary Results and Supplementary File 1). Note that McQ is compatible in principle with any up-stream extraction protocol.

**Fig. 1.**
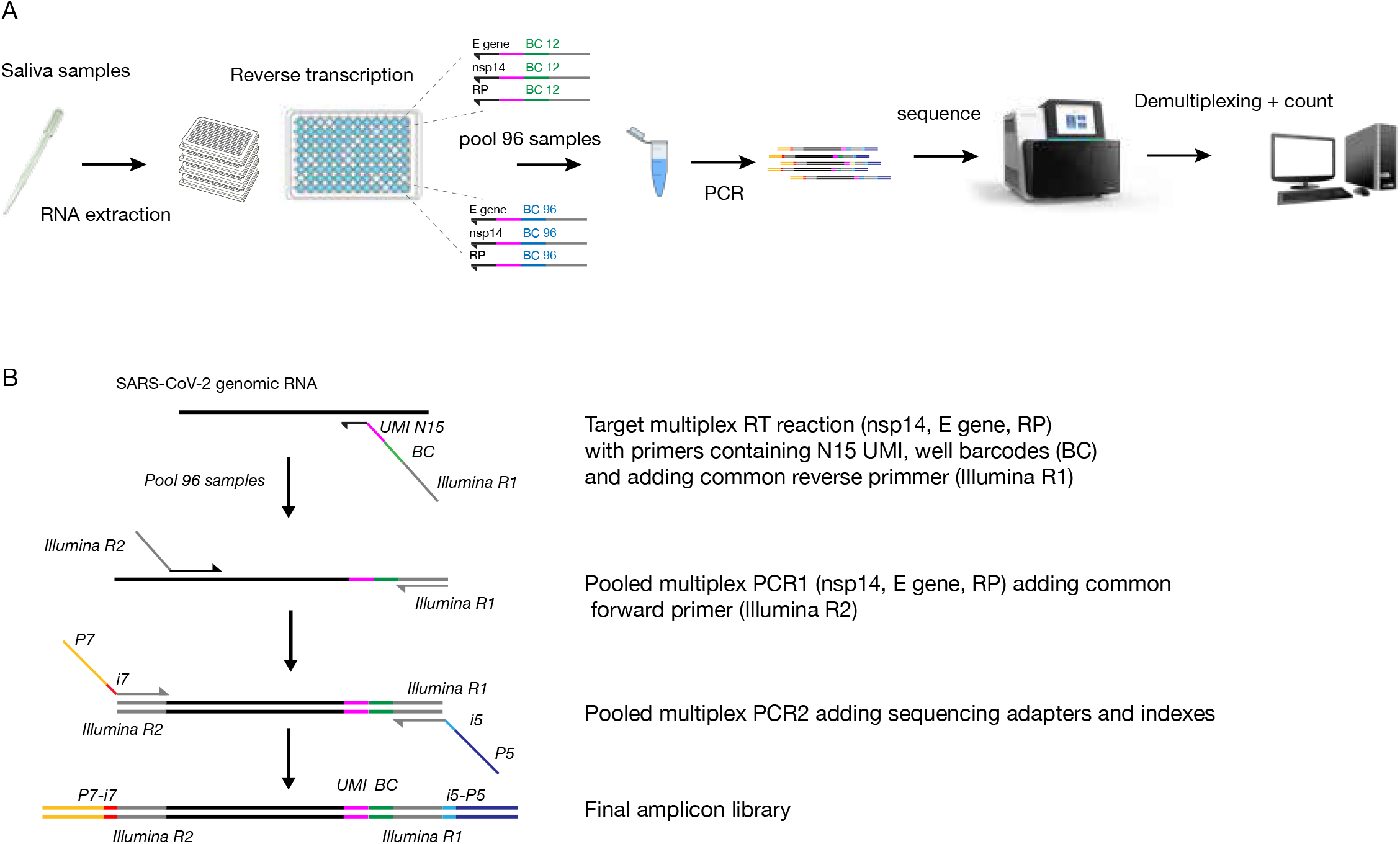
The McQ COVID-19 diagnostic assay. (A) Schematic showing the workflow of reverse transcription with target-specific barcoded primers, PCR amplification of pooled cDNA, addition of sequencing adapters, Illumina sequencing and computational demultiplexing to quantify viral loads. (B) Multiplexing strategy. McQ uses target-specific reverse transcription (RT) primers to introduce a sample barcode (BC, green) during RT, enabling pooling of 96 samples for PCR enrichment of target sites. RT primers contain a unique molecular identifier (UMI) to label unique RNA templates in the sample, and a partial Illumina R1 primer to serve as common reverse primer during PCR1. Viral E and nsp14 as well as human RP regions are reverse transcribed and amplified in multiplexed reactions within a sample. The pooled cDNA of 96 samples is used as template in the first PCR to enrich target sites using three target-specific forward primers and the common reverse primer. Indexed P5 and P7 sequencing adapters are added in a short final PCR for sequencing of multiple 96-well plates in one run. The final amplicons contain three types of barcodes: UMIs to assign reads to unique RNA templates, sample barcodes (BC) to assign reads to a well of a 96-well plate, and i5/i7 indexes to assign reads to a 96-well plate.

The McQ workflow starts with the distribution of extracted RNA into 96-well plates. McQ is designed to quantify the abundance of two viral targets, the E gene and nsp14 (ORF1) regions of the SARS-CoV-2 genome (using primers established by Charite ^30^ and HKU ^31^); and a region of the human RNAse P (RP) gene (recommended by the CDC ^32^). The two SARS-CoV-2 targets serve as internal replicates for viral load. The human target ensures the generation of amplicons even in negative samples to guarantee pool complexity at the level of sample barcodes. The protocol uses a homemade reverse transcriptase as well as DNA polymerase that were benchmarked in the context of this study (see purification protocol in Supplementary File 2 and benchmarking in Supplementary Results).

Reverse transcription (RT) is performed using target-specific primers containing a barcode that uniquely labels each sample. Additionally, the RT primers add a UMI to each cDNA molecule, enabling to identify duplicates that may arise at the following PCR amplification step. After RT, excess primer is digested using Exonuclease I, and equivalent volumes of samples are pooled in batches of 96 for bead purification. The pooled cDNA is used as template in a PCR with three forward primers against individual targets (E, nsp14, RP) and a common reverse primer that anneals to the universal sequence introduced at the RT step. The PCR primers contain overhangs that allow adding indexed sequencing adapters compatible with Illumina sequencing in a second PCR. This enables direct multiplexing of tens of thousands of samples in one sequencing run. Our approach relies on sequences that are compatible with standard Illumina indexing and sequencing (Illumina read1 and 2, and P5 and P7 primers).

Samples are typically sequenced on a MiSeq or NextSeq platform using 75bp single end reads. Samples were demultiplexed using the plate and sample indexes. Several filters were implemented to remove primer dimers and other sources of noise. Then UMIs were counted for SARS-CoV-2 (E and nsp14) and human (RP) targets (Methods). The sample processing protocol is provided in Supplementary File 4. It can be completed within 4 hours, is easily scalable, and cost-efficient at <1 Euro/sample (from RNA to sequencing-ready library). We describe implementation of the full McQ workflow, including RNA extraction and production and integration of homemade enzymes in the Supplementary Results section.

### Establishing McQ assay sensitivity using synthetic SARS-CoV-2 RNA template

To determine the sensitivity of McQ we processed 96 samples spiked with 0 to 100,000 copies of synthetic viral RNA (0-5000 cp/µl RNA sample). Swab or saliva samples carry heterogeneous amounts of human DNA and RNA. To test the effect of this heterogeneity, we added variable amounts of commercial human RNA or RNA/gDNA mix to each sample.

We retrieved all the 96 sample barcodes used to tag these samples, demonstrating the robustness of our approach to sample drop-off during pooled amplification. We observed a good correlation in the UMI counts for the E and nsp14 amplicons, suggesting that both viral primer sets have similar sensitivity. Moreover, this high concordance shows that McQ quantification is not the result of particular biases of a specific primer set. Viral UMI counts were highly similar in presence of various amounts of human DNA and RNA (Figure 2). While the levels of the RP amplicon varied between samples, it did not significantly affect the counts for viral amplicons in the same sample. Together, this suggests that McQ is robust to the heterogeneity in human RNA and DNA content of swab and saliva samples.

**Fig. 2.**
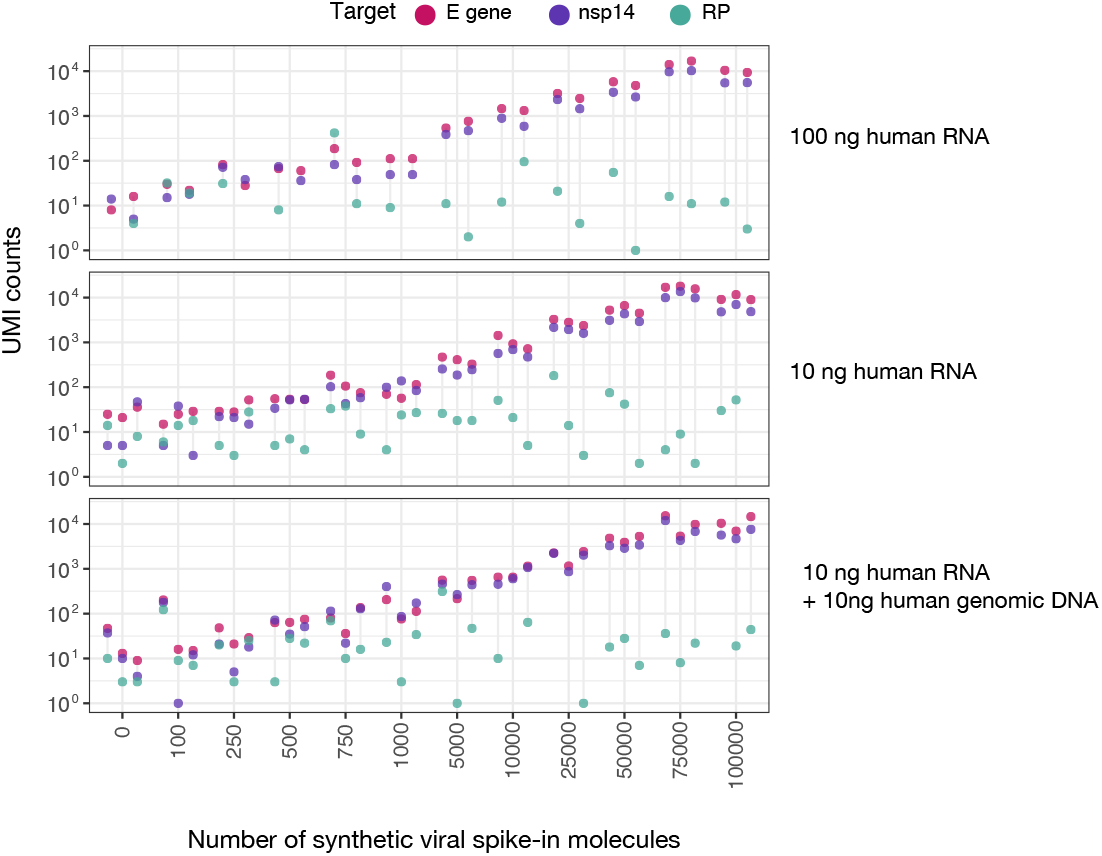
McQ quantitatively detects viral RNA on synthetic templates. 96 samples containing defined numbers of synthetic SARS-CoV-2 RNA template (0-100,000 copies) and human RNA were processed using the McQ workflow. Scatter plots depict the number of Unique Molecular Identifiers (UMI) labeling viral targets (E gene: pink, nsp14: purple) or human control (RP: green) as a function of the number of synthetic viral RNA molecules spiked into total human RNA. Each viral spike-in amount was processed with varying amounts of human RNA mixed into the sample (100 ng RNA, 10 ng RNA, 10 ng RNA + 10 ng genomic DNA) in duplicate or triplicate. UMI counts reflect unique RNA templates present in the sample.

We observed a good correlation between viral UMI counts and the number of SARS-CoV-2 molecules spiked into the sample (Figure 2), and high consistency between replicates, demonstrating that McQ accurately quantifies SARS-CoV-2 RNA. We observed background counts between 4 and 47 viral UMIs in control samples where no SARS-CoV-2 template was present. Similar viral counts were observed for samples containing up to 250 SARS-CoV-2 template molecules (12.5 cp/µl RNA sample), indicating that McQ sensitivity is in the range of 500 copies. This corresponds to 50 copies/µl in the extracted RNA sample.

### Detection of SARS-CoV-2 in clinical samples

Having established McQ robustness and sensitivity, we benchmarked its ability to quantify viral RNA in a panel of 838 clinical samples previously characterised by qPCR in a clinical diagnostics facility. We observed a good concordance between UMI counts for viral targets and Ct values determined by the RT-qPCR assay (Figure 3A). As for synthetic samples we detected all expected barcodes in each pool and consistent UMI counts for E and nsp14 regions. Viral UMI counts showed a linear correlation with Ct values for Ct values below 30, after which the counts flattened out to background levels. This suggests a limit of detection of our assay corresponding to a Ct of around 30 in RT-qPCR. This corresponds to approximately 50 copies/µl (estimates from Robert Koch Institute ^33,34^) and is in line with our results with synthetic RNA (Figure 2). UMI counts for the RP amplicon were relatively constant across samples (Figure 3). We could not detect the RP target in 10% of the samples (85 out of 838). Sample to sample variability in RP counts likely reflects heterogeneity in sampling, extraction and RNA quality.

**Fig. 3.**
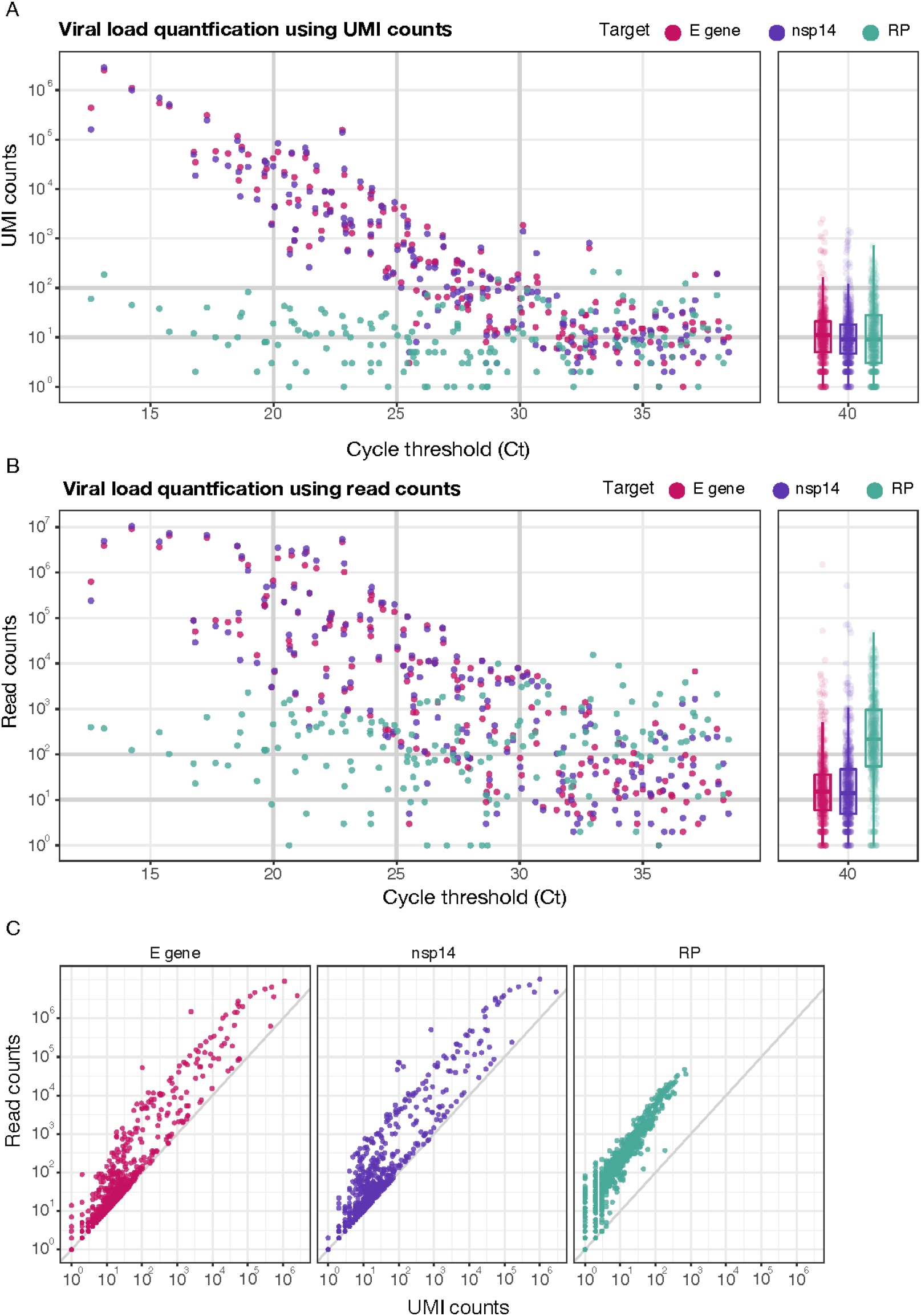
Detection of SARS-Cov-2 RNA in 838 clinical samples using McQ. (A) Scatterplot shows a comparison of UMI counts obtained by McQ and RT-qPCR results on the same sample for 838 clinical samples distributed across 10 plates. Unique Molecular Identifiers (UMI) label viral targets (E gene: pink, nsp14: purple) or human control (RP: green). UMI count distributions for negative samples (Ct 40) are shown as boxplots. (B) Scatterplot depicting the number of reads for each target compared to the qPCR results obtained on the same sample. Read count distributions for negative samples (Ct 40) are shown as boxplots. (C) Plots show a comparison of UMI and read counts for E (pink), nsp14 (purple) and RP (green) targets in the 838 samples from A and B.

Compared to our experiment with synthetic control samples, we observed higher background UMI counts in clinical samples previously diagnosed as negative (Ct of 40) by RT-qPCR (Figure 3). Samples with Ct 40 had acceptable median UMI counts of 9 and 11 for the nsp14 and E targets respectively. However, the background UMI counts reached up to 2452 for some samples, that is two orders of magnitude higher than the maximum of 47 observed in synthetic samples. The high background not only caused false positive sample assignment but also prevented distinction of samples with low viral load from negative samples, limiting sensitivity. We suspect that the presence of many samples with high viral load on these plates, combined with repeated usage of these samples has increased the chance of sample cross-contamination and is a major source of high background. Importantly, this issue was resolved when repeating the procedure on freshly collected samples (Figure 4). Moreover, in the Supplementary Results section we show that inclusion of an Exonuclease I step prior to sequencing lowered viral UMI counts in control samples.

**Fig. 4.**
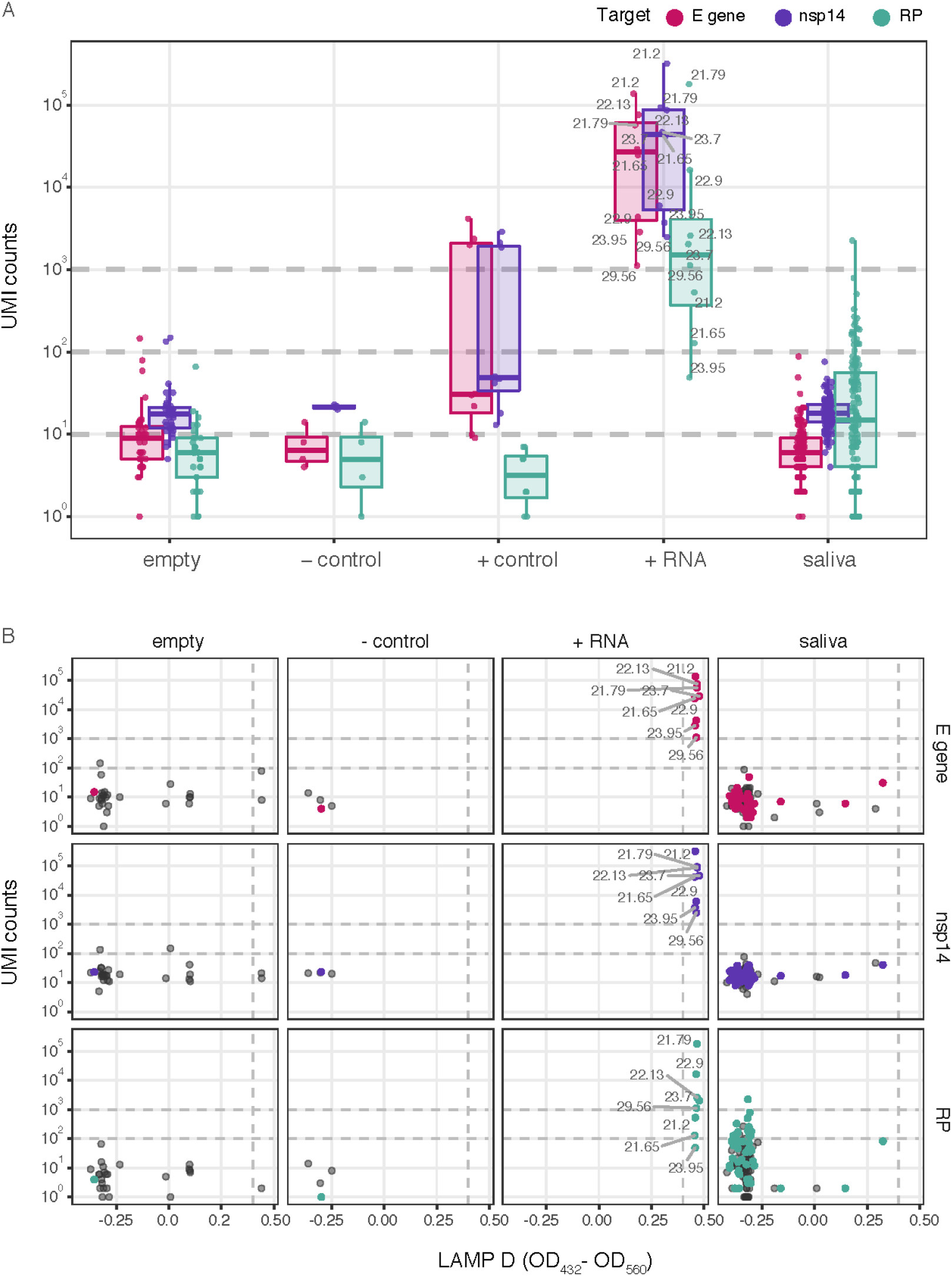
Detection of SARS-CoV-2 RNA in extracted saliva samples. (A) Plots show the number of UMI detected for viral targets (E gene: pink, nsp14: purple) or human control (RP: green) in RNA extracted from 136 saliva samples provided by healthy volunteers (saliva). Empty = water, added before RNA extraction, negative control = saliva from a previously tested negative sample, positive control = highly diluted negative saliva sample spiked with RNA from a sample tested positive for SARS-CoV-2 and 100,000 molecules of Twist synthetic RNA (in water, spiked in prior to extraction), positive RNA = extracted RNA from samples tested positive for SARS-CoV-2 in a clinical diagnostics facility (Ct values indicated in dark grey), added to assay plates after RNA extraction. Boxplots are overlaid to visualize distributions. (B) Comparison of UMI counts for E (pink), nsp14 (purple) and RP (green) targets with D values obtained from a LAMP assay on the same samples. Black dots denote failed LAMP assays (D IC < 0.4).

A unique feature of the McQ protocol is the introduction of UMIs to correct for PCR duplicates in target quantification. We evaluated the importance of this feature by comparing the accuracy of target quantification using simple read counts (Figure 3B) and upon UMI correction (Figure 3A). While the linear correlation between Ct and counts for viral targets was observable with both metrics, the data were substantially more noisy using read counts only (Figure 3B,C). Moreover, background counts in negative samples were significantly higher with up to 1.5 million counts (Figure 3B). We conclude that UMI-based quantification is important for higher accuracy in quantification of viral load by correcting for amplification bias. These results demonstrate that McQ is able to identify positive samples and UMI counts quantitatively reflect viral load.

### Proof of concept of McQ in routine testing

We performed a small pilot study to demonstrate the applicability of McQ for surveillance testing. Saliva samples were self-collected by 136 healthy volunteers, registered online and dropped off at a central collection point. We performed automated RNA extraction from these samples (Supplementary File 1) and applied the McQ analysis framework. In parallel, we processed the same RNA samples with a commercially available loop-mediated isothermal amplification (LAMP) rapid colorimetric assay kit. We included several negative (water and previously tested negative swab samples) and positive controls (100,000 copies of synthetic RNA, dilutions of titrated swab samples, RNA of positive samples). RNA extraction time on the robot was about one hour per 96 well plate. The complete turnaround time was 48h. With these fresh samples, viral UMI counts for negative controls were low (between 4 and 23 counts). This argues for low levels of cross-contamination inherent in our procedure and suggests the high levels of background we observed before were a consequence of contamination already present in the RNA samples. Importantly, the viral UMI counts in positive controls were more than an order of magnitude higher than in negative controls and we could detect positive control samples with low viral load (Ct 29.56) (Figure 4A). We diagnosed all of the saliva samples as negative based on the abundance of E and nsp14 UMIs, which were at comparable levels to negative controls (1-88 UMIs). The results were confirmed independently using LAMP (Figure 4B), yielding 100% agreement between the two approaches. Negative status of three samples with McQ or LAMP values just below the positive threshold was confirmed via qPCR.

As McQ is based on sequencing, a significant part of the costs per sample is coming from Illumina sequencing. To address the impact of sequencing depth on diagnostic accuracy we subsampled the data. With 2600 reads per sample we detected either the E or nsp14 target in all sixteen positive controls, and 15/16 with 260 reads per sample (Figure S10). This suggests that a sequencing depth of 3000 reads per sample could be sufficient to detect positive samples. Together these data demonstrate that McQ has the sensitivity and the throughput to test for presence of SARS-CoV-2 in saliva or swab samples.

## Discussion

We have developed McQ, a SARS-CoV-2 quantification assay that couples early-stage barcoding with next-generation sequencing for multiplexed processing of thousands of samples at low cost, enabling testing at the scales required for routine surveillance screening. Barcoding at the reverse transcription step enables early sample pooling to reduce hands-on time and cost. Integration of homemade enzymes and reagents circumvents potential bottlenecks due to reagent shortages and facilitates implementation of the workflow in settings with limited financial resources. We have demonstrated that McQ can detect SARS-CoV-2 RNA in extracted RNA from clinical samples. Compared with read-based quantification, the UMI-based quantification implemented in McQ enables higher accuracy in determination of viral load by correcting for amplification bias. Viral UMI counts were predictive of Ct values obtained on the same samples using a validated RT-qPCR method in a clinical diagnostic facility. Moreover, McQ accurately diagnosed samples in an integrated testing workflow entailing self-sampling and automated RNA extraction from saliva. McQ is highly modular and can easily be extended to other human or SARS-CoV-2 targets, or to multiplexed detection of several pathogens in one sample.

McQ analytical sensitivity is lower than with gold standard RT-qPCR based tests at 50 copies/µl, corresponding to samples with a Ct of approximately 30. Nevertheless, this may be sufficient for efficiently limiting viral spread as epidemiological studies have identified high testing frequency and fast turnaround times as the most critical variables for controlling transmission ^35,36^. Sensitivity was only secondary to these factors, with little difference in preventing transmission with high (10^3^ cp/ml, RT-PCR) or lower sensitivity (10^5^ cp/ml, e.g. LAMP) tests ^35^. In addition, samples with low viral load (Ct >30 or <50 copies/µl RNA) have been associated with a loss of cultivation of the virus ^33,34,37–40^ and could pose only a low risk for transmission. Combining McQ with our in-house RNA extraction workflow from saliva we were able to clearly distinguish positive from negative samples, with viral UMI counts at least an order of magnitude higher in positive samples even for a sample with low viral load (Ct 29.56). McQ increases testing capacity at low cost and could complement highly sensitive symptomatic testing, which on its own is not sufficient to control viral spread due to high infectivity of pre- and asymptomatic individuals ^5,7^.

McQ is based on homemade reverse transcriptase and polymerase enzymes. We provide detailed purification protocols and recipes for non-proprietary buffers for use of our enzymes by the community. Our homemade MMLV_mut reverse transcriptase and Pfu-Sso7d polymerase enzymes showed comparable performance to commercial enzymes. Our in-house solutions lower reagent costs and circumvent the dependence on commercial kits that are subject to shortage when highly demanded. This will facilitate the implementation of widespread testing. Furthermore, they can be readily deployed in other testing frameworks and complement open-source solutions for one- and two-step RT-qPCR diagnostics ^41–44^.

Reducing molecular contamination, shortening turnaround time, and investing in alternative methods for swab or saliva sample processing that are more automation friendly are key areas for improvement towards implementation of McQ in a surveillance testing framework. In the first set of RNA samples we processed with McQ we detected high numbers of viral UMIs in a significant number of negative (Ct 40) samples, causing false positive sample assignment and limiting sensitivity by preventing distinction of samples with low viral load from negative samples. With freshly collected samples viral UMI counts for negative controls and in negative saliva samples were reduced significantly, suggesting that the high levels of background we observed before were a consequence of contamination already present in these RNA samples. Moreover, we have shown that inclusion of an Exonuclease I step prior to sequencing reduces viral UMI counts in control samples. Combining this improvement with strategies to reduce molecular contamination (according to clinical testing guidelines) should further reduce background levels of viral UMI counts. These measures will improve sensitivity, and reduce the potential for false positives that require re-testing.

### Supplemental results

#### Automated paramagnetic bead-based RNA extraction from swabs and saliva

RNA purification is a major bottleneck in scaling up testing capacity. To overcome this we implemented the SARS-CoV-2 RNA purification protocol from BOMB-Bio ^45^ on the Biomek i7 liquid-handling platform at the EMBL Genomics Core Facility. A detailed description of the automation workflow, reagents and the source code for sample processing on the Biomek i7 platform is provided in Supplementary File 1.

We first implemented bead-based extraction manually using swab samples. Nose and throat swab samples were collected in Amies transport medium from COVID-19 patients at Uni Klinikum Heidelberg, Frankfurt and Oldenburg (Methods). 200 µl of each sample was manually transferred to a 1.2 ml deep-well plate and 100 µl GITC lysis buffer was added. Samples were subsequently purified using 40 µl of paramagnetic beads, washed with ethanol and resuspended in 25 µl of MilliQ water. To evaluate extraction efficiency we performed RT-qPCR (using TaqMan Fast Virus 1-Step Master Mix, Thermo) on a subset of extracted samples (Figure S1). These included extractions of Expi293F cell suspensions (10^6^ cells/ml) in a 0.9% NaCl saline solution with different amounts of synthetic SARS-CoV-2 RNA template spiked-in (2.5 – 500 copies/µl corresponding to 500-100,000 copies total), dilution series of two swab samples previously tested positive for SARS-CoV-2 by RT-qPCR (swab 1 Ct = 16.37, swab 2 Ct = 16.61), and an undiluted SARS-CoV-2 negative swab sample (swab 3). We obtained similar Ct values for identical amounts of synthetic viral template regardless of whether it was spiked in prior to extraction or immediately prior to RT, indicating high extraction efficiency (Figure S1A). Bioanalyzer traces confirmed high RNA quality (Figure S1B). We detected viral RNA by qPCR in all swab dilutions (Figure S1C) and observed an increase in Ct with increasing dilution, as expected. RNA quality was generally lower in extracted swab samples (Figure S1D) compared to human cell control or synthetic viral RNA spiked into Expi293F cells. The differences in quality could be due to repeated freeze-thawing of swab samples, or inhibitory effects of the swab collection medium during RNA extraction. Having demonstrated that bead-based RNA extraction is highly efficient from manually processed swab samples, we proceeded to implement automated extraction. At this point we turned to saliva samples as sample collection is easier and can be performed by individuals, without the need for trained personnel. Saliva samples provided by healthy volunteers were collected in a 0.9% NaCl solution. Samples were inactivated in bulk by the addition of boiling water previously heated to 95°C to a 1L beaker containing all sample tubes, and incubated for 5 minutes. Water temperature was required to be >80°C after the 5 min incubation. After inactivation, 200 µl of each sample was manually transferred to a 1.2 ml deep-well plate and 100 µl GITC lysis buffer was added. Samples were subsequently purified using 40 µl of paramagnetic beads, washed with ethanol and resuspended in 25 µl of MilliQ water. RNA quality was similar to manually extracted swab samples (Figure S1E).

The complete extraction procedure can be achieved in 145-175 min /plate (∼40 min for sample collection, boiling and scanning of barcodes; 60-90 min for manual transfer to 96-well plates; 45 min for robotic extraction). Automated extraction typically yielded 10 - 75 ng of RNA for saliva samples that were subsequently used for reverse transcription.

### Expression and purification of homemade enzymes

To develop a workflow that is independent of supply-limited and expensive commercial enzymes and reagents we expressed and purified a Moloney Murine Leukemia Virus-based reverse transcriptase (MMLV_mut) and a Pfu-Sso7d polymerase (described in detail in Supplementary File 2). Both enzymes were purified using a multi-step purification to obtain highly pure MMLV_mut (76 kDa) and Pfu-Sso7d (98 kDA) protein. MMLV_mut was expressed in E. coli BL21 (DE3) cells, with a yield of 120 mg MMLV_mut from two litres of expression culture. Pfu-Sso7d was expressed in E. coli Rosetta (DE3) cells, with a yield of 17 mg Pfu-Sso7d from one litre of expression culture.

### Optimising MMLV_mut reaction conditions

To establish reaction conditions for homemade MMLV_mut reverse transcriptase we performed cDNA synthesis from synthetic SARS-CoV-2 viral RNA template with the N1 primer set recommended by the CDC, and evaluated efficiency via qPCR using SYBR Green master mix (Table S1, Figure S2A). In a first attempt to test its activity we used MMLV_mut at the suggested concentration of commercial SSIII (1 µl of 1 mg/ml stock in 20 µl reaction) with a buffer similar in composition to commercial SSIII buffer (Buffer V1). Following recommended reaction conditions for SSIII we obtained cycle threshold (Ct) values of 28 and 31 (for 100,000 and 10,000 spike-in molecules) after reverse transcription with MMLV_mut, outperforming the commercial SSIII enzyme (Ct 30 and undetermined, respectively) and demonstrating MMLV_mut was active and produced cDNA suitable for SARS-CoV-2 detection. To identify robust conditions for efficient reverse transcription using homemade MMLV_mut enzyme we extensively varied different reaction parameters (Figure S2A). We swapped the target-specific N1 reverse transcription primer for random hexamers to avoid optimizing conditions for a specific target. We found a Tris and MOPS based buffer with reduced salt concentrations and glycerol (for thermostabilisation) and a non-ionic detergent resulted in robust reverse transcription (Buffer V5, developed by Alexander Klenov ^46^). Reverse transcription with 10x less enzyme (stock of 0.1 mg/ml) substantially improved efficiency (Ct 24.76 compared to Ct 31 with 1 mg/ml enzyme stock for 10,000 spike-in molecules). Further dilution of the enzyme stock (to 0.02 mg/ml) tended to result in decreased Ct but with considerably more variable outcomes (Figure S2A). We applied the optimized conditions (0.02 mg/ml enzyme and V5 buffer) to determine SARS-CoV-2 load in four previously tested clinical samples (kindly provided by M. Knop, ZMBH + DKFZ) and obtained very similar Ct values (Fig. S2B). The previous RT-qPCR assay was run in a clinical diagnostics facility, using a commercially available test kit that uses altered versions of the E Sarbeco ^30^ primer set and using 10 µl RNA as input, which could explain the small discrepancies. Taken together we identified robust reaction conditions for homemade MMLV_mut reverse transcriptase compatible with efficient production of cDNA, yielding similar viral load quantification as a validated qPCR kit.

### McQ amplicon and primer design

We designed primers targeting the E Sarbeco and nsp14 (ORF1) regions in the SARS-CoV-2 genome for next-generation sequencing based multiplexed detection. These regions were used in RT-PCR tests developed by the Charite ^30^ and HKU ^31^, and both sets of primers resulted in high analytical efficiency and sensitivity ^47,48^. We modified priming sequences to increase melting temperatures and ensured specificity via blast ^49^ against the human genome (hg38). To ensure primers are uniquely targeting and capable to amplify SARS-CoV-2 variants we aligned primers to a reference SARS-CoV-2 genome (NC_045512.2) and a panel of 1775 sequenced SARS-CoV-2 strains obtained from GISAID. We based RP primer design on the sequences recommended by the CDC, for amplification of a human control target, and only modified primers to match melting temperatures of viral primers. It is important to note that while these primers can be used to ensure pool complexity at the level of sample barcodes, they cannot distinguish negative samples from failed extractions as human genomic DNA contamination would lead to the presence of RP amplicons ^50^. Designing the RP reverse transcription primer to target another exon will address this problem. All primers were additionally checked for (self)-dimerization potential. To barcode samples at the cDNA step we incorporated 15 nucleotide sample barcodes (described in Hossain et al. ^25^) in reverse transcription primers, to allow pooling of 96 cDNA samples into a single tube for PCR amplification. Reverse transcription primers additionally contained a N15 UMI (for viral load quantification) and a partial Illumina read1 primer sequence. This sequence serves as a common reverse primer for all three targets in the first round of PCR amplification, and is used in the second PCR to add indexed sequencing adapters. PCR1 forward primers were correspondingly designed with a partial Illumina read 2 primer sequence in their overhangs. This setup allows the direct sequencing of amplicons and readout of sample and sequencing indexes using standard Illumina reagents and kits. Samples are sequenced in 75SE mode and demultiplexed into 96-well plates using 8nt i5 and i7 indexes. The first 15 cycles of read 1 contain the sample barcode to assign samples to plate wells, followed by 15 cycles of UMI that identify unique RNA template molecules and allow correcting for amplification bias during PCR, the target-specific priming sequence (18 to 24 nt) and 21 to 27 nt of target sequence (depending on the target) that allows distinguishing between actual reads and primer dimer. The 27 nt of human RP sequence are too short and do not contain sufficient genomic variation to identify individuals from sequencing data.

### Implementation of McQ, an NGS-based assay for multiplexed SARS-CoV-2 detection

For initial protocol implementation we used 1 µl of the 1 mg/ml MMLV_mut stock with buffer V1 and performed reverse transcription of defined mixtures of synthetic SARS-CoV-2 viral RNA and synthetic human liver RNA as template. To maximize Pfu-Sso7d amplification efficiency we tested amplification of the E amplicon from cDNA generated from 5000 copies of synthetic viral spike-in with four reaction buffers (HF-V2 to HF-V5, described by Alexander Klenov) ^51^ in addition to HF-V1, our customary in-house Pfu-Sso7d buffer. Two buffers, HF-V4 and HF-V5 enabled strong and specific amplification at levels similar to commercial Phusion enzyme (Figure S3A). In contrast to other buffers HF-V4 and HF-V5 contained Arginine and Trehalose, which could aid the amplification of low amounts of template. We proceeded with buffer HF-V5, which enabled reduced primer dimer formation compared to HF-V4, as evidenced by the substantially weaker primer dimer band. We determined optimal primer concentrations and viral to human primer ratios during reverse transcription and PCR1 to minimise the formation of primer dimers and favour amplification of viral targets, settling on a 2:1 ratio of viral over RP primers in both reverse transcription (200 nM and 100 nM) and PCR1 (50 nM and 25 nM). Sequencing adapters were added in PCR2 using commercial KAPA polymerase. We applied the resulting protocol to twelve samples containing defined amounts of synthetic SARS-CoV-2 spike-in molecules (10 to 100,000) mixed with 10 ng total human RNA. To address whether pooled amplification leads to sample dropout or reduces quantification accuracy we performed both PCRs on individual samples (Figure S4A) and on pooled cDNA of the twelve samples (Figure S4B), using 35 and 30 cycles in PCR1, respectively. We obtained similar UMI counts (labelling unique RNA templates) for E and nsp14 primer sets within a sample, indicating comparable sensitivity, and UMI counts between replicates showed good agreement. Viral UMI counts showed a linear increase with the amount of synthetic SARS-CoV-2 template added to samples. UMI counts for the RP amplicon were relatively similar across samples, as expected from the fixed amount of human RNA spiked into each sample. We detected all twelve sample barcodes in individually amplified samples, as well as in the sample pool. While the total UMI counts for a given template amount were lower in the sample pool than in individually amplified samples, the linear, positive correlation of viral UMI counts with the amount of synthetic SARS-CoV-2 template added to samples was preserved (Figure S4B).

We next replaced commercial KAPA with Pfu-Sso7d in PCR2 and identified optimal buffer and annealing conditions (Figure S3B). Using cDNA from the pool of twelve samples described above we performed PCR1 with Pfu-Sso7d and compared performance of KAPA and Pfu-Sso7d in PCR2. Both polymerases yielded similar numbers of UMIs (Figure S5A). KAPA polymerase is favored in library applications due to its low GC-amplification bias compared to other polymerases. We quantified amplification bias by counting the numbers of reads per UMI and found very similar distributions for Pfu-Sso7d and KAPA, with a tendency for less variation in read numbers with Pfu-Sso7d (Figure S5B). We could not compare KAPA polymerase against Pfu-Sso7d for PCR1 as KAPA consistently produced non-specific higher molecular weight bands for the RP and nsp14 targets. As PCR1 entails more cycles than PCR2 it is possible that a larger difference in efficiency and bias between the two polymerases is masked when comparing them in the second PCR only.

### Sources of and strategies to improve incomplete UMI recovery

E and nsp14 UMI counts in samples with synthetic SARS-CoV-2 RNA were 5 to 10-fold lower than expected based on the number of viral template added to these samples (Figure S4, S5). Individually amplified samples showed the same tendency, particularly for samples with high amounts of template added (Figure S4A). This indicates that sample pooling was not by itself the cause for reduced UMI recovery. Subsampling of reads revealed that higher sequencing depth only incrementally increased UMI numbers. Taking into account the moderate amplification bias (Figure S5B) this led us to believe that steps during library preparation, such as inefficient or biased reverse transcription and the first step of PCR, presented bottlenecks for UMI recovery. We first addressed the impact of optimizing reverse transcription. As for random hexamers, buffer V5 and a 10-fold dilution of MMLV_mut (0.1 mg/ml stock) improved reverse transcription efficiency with McQ target-specific viral primers (Figure S6A), demonstrated by the higher number of viral UMIs detected with the diluted enzyme in buffer V5. Improved viral UMI recovery with 0.1 mg/ml compared to 1 mg/ml MMLV_mut was more pronounced in buffer V1 than V5, suggesting that multiple interacting factors can lead to improved efficiency. The improvement in efficiency was target specific as UMI counts for RP dropped with lower enzyme concentration in both buffers. After a series of additional experiments we converged on an optimal MMLV_mut-based reverse transcription protocol using buffer V5^46^ with trehalose (buffer V7) and 0.1 mg/ml MMLV_mut (see Figure S6C for comparison to buffer V1 and 1 mg/ml MMLV_mut in RT-qPCR). We decided against using a further dilution of MMLV_mut (to 0.02 mg/ml stock) due to the higher variability in results (Figure S2A). We also compared MMLV_mut to commercial SSIV enzyme (Thermo), which promises enhanced yield and performance, at different concentrations and incubation temperatures but did not observe any improvement in UMI recovery for viral targets with SSIV (Figure S6B). The RP target was amplified better with SSIV in all tested conditions.

PCR jackpotting (the selection of templates in the first round of PCR) is another well-known bottleneck in library preparation. To estimate the relative contribution of bottlenecks in reverse transcription and PCR on reduced UMI recovery, and address whether a split amplification design aimed at randomizing effects of PCR jackpotting would improve UMI recovery we amplified two pools of cDNA, each consisting of twelve samples, using four different PCR designs (Figure S7A). Each PCR design used the same pool of cDNA as input. The twelve samples contained defined amounts of synthetic SARS-CoV-2 spike-in molecules (0 to 100,000, in duplicate) mixed with 10 ng total human RNA. For pool 1 we performed reverse transcription using a modified buffer V5 and 0.1 mg/ml MMLV_mut (optimized conditions) while pool 2 samples were reverse transcribed using the initial conditions (buffer V1 and 1 mg/ml MMLV_mut). As the cDNA input in all four designs was the same, calculating the fraction of UMIs that are shared between the resulting samples can address whether PCR substantially biases the subset of UMIs amplified from a cDNA pool (Figure S7B). We observed reasonable overlap (44%-57%) between samples derived from the same PCR1 reaction (AA and AB, as well as BA and BB), but almost no overlap between samples deriving from separate PCR1 reactions (AA or AB to BA and BB), indicating almost entirely distinct subsets of UMIs were amplified in each PCR1. This strongly suggests PCR jackpotting as a major source limiting UMI recovery. To address whether the number of UMIs could be improved by splitting a single PCR into four reactions, randomizing PCR jackpotting, we compared the number of UMIs recovered with each design (Figure S7C). Interestingly, we observed no difference between designs for the pool of samples reverse transcribed with less optimal conditions (pool 2), but improved UMI recovery with designs using PCR splitting in one or both PCRs in the pool of samples reverse transcribed with optimized conditions (pool 1). No UMIs should be shared between pools 1 and 2 as they result from distinct reverse transcription reactions and UMI complexity is sufficiently high (N15). UMIs that are shared between pools 1 and 2 (1%-6%) therefore represent the level of contamination accrued from molecular contamination at any step after reverse transcription or from sample mis-assignment. Based on these results we suggest further optimisation of the protocol towards higher UMI recovery should address the PCR step. The split amplification design we used here can be readily implemented without further experimental optimisation, with the disadvantage of an increase in PCR sample numbers. Alternatively, evaluation of other polymerases or buffer conditions that ameliorate jackpotting, such as adding Betaine or prolonging initial denaturation ^52^ for Pfu-Sso7d could help.

### Sources of and strategies to improve background signal

Reads mapping to viral sequences in samples containing no SARS-CoV-2 RNA are a source of false positive test outcomes and limit sensitivity of detection assays. Non-zero background has been observed in other sequencing-based assays for SARS-CoV-2 detection ^23,24,53,^ and molecular contamination and mis-assignment of sequencing reads were identified as main underlying sources ^23^. Leveraging the UMI information we have shown that between 1% and 6% of the UMI counts in a pool are contaminants of another pool (Figure S7B), resulting from events taking place after reverse transcription (during library amplification or sequencing). Index hopping during sequencing due to free index primers that were not used up in PCR is a major source for sample misassignment ^54–56^. To specifically address contribution of this variable to the fraction of NTC UMIs (viral UMIs in samples without SARS-CoV-2) we compared the number of viral UMIs in 63 NTC samples prior to and after an additional Exonuclease I digestion step after PCR2 that removes excess index primer. The NTC samples were part of pools of 12 samples that contained 2 to 12 NTC samples. Exonuclease I treatment reduced NTC UMIs at least partially in 72% of samples, and completely eradicated NTC UMIs in 58% of samples (Figure S8). The extent of reduction was pool specific suggesting that molecular contamination prior to the reverse transcription step varied between pools. In 25% of samples the number of NTC UMIs increased, indicating additional sources of NTC UMIs during sequencing (e.g. contamination in the sequencers). These data suggest that index hopping during sequencing resulting from free-floating indexed primers accounts for a substantial fraction of background that can be reduced by including an Exonuclease I treatment step after PCR2 and prior to sequencing.

### Automation of the McQ reverse transcription step

To further reduce hands-on time in the McQ workflow we implemented a protocol for automated reverse transcription on the Beckman i7 automation system at the EMBL Genomics Core Facility. The method can easily be implemented on any liquid-handling system, using the original manual protocol as a guideline. A detailed description and step-by-step protocol is provided in Supplementary File 3. To compare performance to manually processed samples we performed reverse transcription using 10 µl aliquots of the same 96 samples (containing varying levels of synthetic viral RNA spike-in and human input material) used for Figure 2. As for manually processed samples, we detected reads for all 96 sample barcodes (Figure S9A), and observed very good concordance between UMI counts from manually processed samples and samples processed on the i7 (Figure S9A, B). The protocol is currently implemented for parallel processing of two plates and takes approximately 106 min (20 min handling time and 86 min off-deck incubation time). It can easily be scaled as all incubation steps are performed off-deck on thermal cyclers.

## Methods

Detailed methods for automated RNA extraction are provided in Supplementary File 1, for production of homemade enzymes in Supplementary File 2 and for robotic implementation of the McQ reverse transcription step in Supplementary File 3. Supplementary File 4 provides a detailed step-by-step protocol for the entire McQ workflow (from extracted RNA to sequencing-ready library). All oligo sequences are listed in Supplementary Tables 1 and 2 and have been deposited at http://github.com/gtca/McQ.

### Samples used in the study

To benchmark McQ we used pseudo-anonymized surplus RNA sample material that had been collected for clinical diagnosis of SARS-CoV-2 infection by RT-qPCR carried out by the diagnostic laboratory of Heidelberg University Hospital. Such reuse of material is in accordance with German regulations, which allow development and improvement of diagnostic assays using patient samples collected specifically to perform the testing in question. Pharyngeal swab specimens provided to us were either collected through the nose (nasopharyngeal) or the mouth (oropharyngeal), or sometimes one swab was used to collect both.

### Clinical sample handling

Specimens were collected as nasopharyngeal and oropharyngeal flocked swabs in Amies medium (eSwab, Copan Italia). The sample collection happened as part of the routine operation of Heidelberg University Hospital and at public testing stations set up by the City of Heidelberg. Collected samples were transported in sterile containers, delivered to the diagnostic laboratory within a few hours, and then examined directly or stored at 4°C until further processing. Samples were processed in a biosafety level 2 cabinet until inactivation by heat or mixing with a lysis buffer.

### RNA isolation and RT-qPCR diagnostics for clinical samples

The standard diagnostic pipeline of the hospital laboratory was as follows: RNA was isolated from nasopharyngeal and oropharyngeal swab specimens using QIAGEN kits (QIAGEN, Hilden, Germany); either automated on the QIASymphony (DSP Virus/Pathogen Mini Kits) or QIAcube (QIAamp Viral RNA Mini Kits) devices or manually (QIAamp Viral RNA Mini Kits). Please note that the QiaCube uses a sample volume of 140 µl and an elution volume of 100 µl, whereas the QiaSymphony uses a sample volume of 200 µl and an elution volume of 115 µl. RT-qPCR for the quantification of the SARS-CoV-2 viral genome was performed using kits and reagents from TIB MOLBIO Syntheselabor, Berlin, Germany. The kits were used according to the manufacturer’s instruction and contained the primer/probe sets developed based on the published Sarbeco primer set ^30^. Per 20-µl reaction, the master mix contained 5.4 µl of RNAse free water, 4.0 µl of LightCycler Multiplex RNA Virus Master (Roche, Basel, Switzerland), 0.5 µl of LightMix Modular SARS and Wuhan CoV Egene (cat. no. 53-0776-96; TIB MOLBIOL Syntheselabor GmbH, Berlin, Germany) or LightMix Modular SARS and Wuhan CoV N gene (cat. no. 53-0775-96; TIB MOLBIOL), 0.5 µl of LightMix Modular EAV RNA Extraction Control (cat. no. 66-0909-96; TIB MOLBIOL), and 0.1 µl of reverse transcriptase enzyme (LightCycler Multiplex RNA Virus Master, Roche, Basel, Switzerland). The master mix (10 µl) was distributed per reaction into 96-well plates, and 10 µl of purified RNA was added per well. The performance of the RT-qPCR was validated using a positive control for the E gene. A total of 1000 molecules of E gene RNA per RT-qPCR reaction correspond to a CT 30.

### Optimisation of MMLV_mut reaction conditions using the N1 primer set

To test homemade MMLV_mut synthetic RNA template (Twist Biosciences, spike-in 1 or 2) was used in the indicated amounts. To each reverse transcription (RT) reaction we added 1 µl 10mM dNTPs, 1µl 2µM gene-specific primer or 50 ng random hexamers (Thermo) in a total volume of 13µl. Following a 5 minute incubation at 65°C reactions were placed on ice.

The indicated homemade RT buffer, 1µl 10mM DTT and 1 µl MMLV_mut (at the indicated stock concentration, diluted from 2 mg/ml stock using MMLV_mut dilution buffer) were added as a master mix and cDNA synthesis was performed at 55°C (for target specific RT primer) or 50°C (for random hexamers) for 30 to 60 min (see Table S1) followed by 15 min inactivation at 70°C. Buffer recipes are provided in Supplementary File 2. For determination of RT efficiency, SYBR green PCR master mix (Thermo) was used according to the manufacturer’s protocol with primers at 100 nM. Nuclease free water was used as a non-template control. Reactions were run on an ABI flex with default SYBR green cycling conditions.

### McQ reverse transcription, cleanup and PCR amplification

A detailed step-by-step protocol is provided in Supplementary File 4. 10 µl of sample (either 10 µl extracted RNA or a mixture of indicated amounts of Twist synthetic viral RNA and total human RNA filled up with nuclease-free water) was used as input to reverse transcription. Mixtures of the three indexed primers and dNTPs were prepared in a 96-well format index master plate as described in Supplementary File 4. The final primer concentrations in reverse transcription reactions were 200 nM for E and nsp14, and 100 nM for RP. Primers, dNTPs and template were incubated at 65°C for 5 min, and immediately placed on ice to cool down for 1 min prior to addition of the enzyme master mix containing MMLV_mut at the indicated concentration (1 µl of indicated stock in a total of 20 µl reaction), the respective buffer, DTT, and RNAse inhibitor (Takara Bio). Reverse transcription was run in a thermocycler (55°C 30 min, 70°C 10 min). Samples were then incubated with thermolabile Exonuclease I (NEB) (37°C 30 min, 85°C 5 min) to digest excess barcoded RT primer. For manually processed samples, samples were processed in 96-well format using multichannel pipettes. For reverse transcription on the Biomek i7 we prepared the index plate as described in Supplementary File 4. RT enzyme and Exonuclease I master mixes were prepared in four 1.5 ml Lo-bind Eppendorf tubes as described in Supplementary Files 3 and 4. The procedure after ExoI digest was the same for manually and robotically processed samples. All samples from a plate (up to 96) were pooled using a multichannel pipette. 10 µl of each well were transferred into 8-well PCR strips, and combined into 1.5 ml Lo-bind Eppendorf tubes and mixed well. Samples were purified using 1X SPRI beads (Beckman Coulter Life Sciences) according to manufacturer’s instructions, and eluted in a total volume of 30 µl of nuclease-free water (NFW). To avoid sample loss due to elution in a small volume of NFW relative to beads we used only 30 µl of beads for cleanup and added 930 µl of SPRI buffer only (for a total volume of 1X); see Supplementary File 4 for details. 29 µl of the cleaned, indexed cDNA pool was used as template for multiplexed amplification with two virus-specific forward primers (E and nsp14, 50 nM each), human RP-specific forward primer (25 nM), and a common reverse primer binding to the constant sequence added with the RT primer (125 nM). Each pool was amplified for 30 cycles with homemade Pfu-Sso7d (0.07 mg/ml) and buffer HF-V5. The master mix and samples were kept on ice at all times during the sample preparation, and reactions transferred directly to the pre-warmed PCR block (98°C). The following cycling conditions were used: 98°C 30 sec; [98°C 10 sec, 62°C 20 sec, 72°C 20 sec] X 30 cycles; 72°C 5 min. PCR products were then cleaned using 1.8 volumes of SPRI magnetic beads (Beckman Coulter, Life Sciences) according to manufacturer’s instructions, and eluted in 15 µl of nuclease-free water (see Supplementary File 4 for detailed procedure).

Sequencing adapters and sample indexes were added in PCR2, using 3 µl of cleaned PCR1 product as input. Indexed Illumina P5 and P7 primers were used at 400 µM each, using unique i5-i7 index pairs for each 96-well plate pool. Each pool was amplified for 10 cycles with homemade Pfu-Sso7d (0.07 mg/ml) and buffer HF-V5. The master mix and samples were kept on ice at all times during the sample preparation, and samples transferred directly to the pre-warmed PCR block (98°C). The following cycling conditions were used: 98°C 2 min; [98°C 10 sec, 62°C 30 sec, 72°C 15 sec] X 10 cycles; 72°C 2 min. Libraries were cleaned using 1 volume of SPRI magnetic beads (Beckman Coulter, Life Sciences) according to manufacturer’s instructions and eluted in 15 µl of nuclease-free water. Sample concentrations were determined using Qubit high-sensitivity DNA assay, and quality confirmed on an Agilent high-sensitivity Bioanalyzer chip. Samples were pooled equally (according to pool size and concentration) for sequencing. We amplified samples individually for data in Supplementary Figures S3, S4 (upper panel), and S6. Procedure was the same as for 96-well pooled amplification, except that samples were cleaned up individually after RT, eluted in 7 µl NFW, which was used as input for PCR1 with 35 cycles. The entire cleaned PCR1 reaction was used as input for PCR2. We processed samples in pools of 12 for Supplementary Figures S4, S7 and S8. To this end 10 µl of each well was pooled, cleaned with 1X SPRI beads, and eluted in 40 µl of NFW. Half of the cDNA pool was used as template for PCR1, which was run identically as for the 96-well pools. PCR1 products were cleaned using 1.8X SPRI beads and eluted in 12 µl NFW. Half of the cleaned PCR1 product was used as template for PCR2.

### Exonuclease I digest to remove excess indexed P5 and P7 primers

To reduce contamination during sequencing caused by leftover indexed primers, the PCR2 products were optionally treated with thermolabile Exonuclease I (NEB) for 30 min at 37°C followed by 5 min at 85°C.

### SARS-CoV-2 detection in saliva samples

Healthy volunteers collected and registered their own samples and dropped them off at a central collection point. Volunteers were provided with a self-testing kit consisting of a 50 ml falcon tube containing 5 ml of 0.9% saline (0.9% NaCl solution, sterile), a barcoded 2 ml tube with screw lid and a disposable pipette inside a plastic bag. The set further contained an information sheet containing the code of the test set and a link to an instruction video for correct sample collection. Volunteers used the saline solution for deep-throat gargling (30 sec) for sample collection, which was spit back into the Falcon tube. The Falcon tube was shaked vigorously and 1-1.5 ml of the mixture was transferred into the 2 ml tube using the plastic disposable pipette. Volunteers were instructed to collect samples before breakfast or brushing their teeth, on the same morning the RNA was extracted. Volunteers dropped their 2 ml sample tube at the established collection points before 9:30am. Sample inactivation proceeded as described above in a safety Level S2 laboratory. For transfer to 96-well plates, samples were processed in a laminar flow hood. The plate was then sealed with a foil and placed on the robot for RNA extraction. RNA extraction and library preparation were performed as described above, with the following deviations: We included an additional 2.2X SPRI cleanup step after RNA extraction, samples were incubated with ExoI after PCR2 to digest excess indexed primer, reverse transcription was performed with commercial SSIII and we used commercial NEB Phusion ready mix in PCR2. Saliva samples were distributed into two 96-well plates, along with positive and negative controls. Positive controls were added to each plate at the same location, and included a highly diluted negative saliva sample spiked with RNA from a sample tested positive for SARS-CoV-2 (spiked into two wells), 100,000 molecules of Twist synthetic RNA (in water, spiked into two wells prior to extraction) both plotted as positive control, and extracted RNA from four samples tested positive for SARS-CoV-2 (added after RNA extraction) plotted as positive sample. We included two kinds of negative controls: Empty wells (water added before RNA extraction), to measure background contamination, and saliva from a previously tested negative sample (spiked into two wells). An index master mix plate was prepared in a PCR hood after UV exposure from newly ordered oligos and in a room where no SARS-CoV-2 related reagents had been handled. For comparison we processed 1 µl RNA from each sample with the SARS-CoV-2 Rapid Colorimetric LAMP Assay kit (NEB) following manufacturers instructions except that reaction volumes were halved and reactions performed in 96-well format. LAMP delta OD value (D) was calculated as the difference between absorbance at 432 nM and 560 nM (D = OD432 – OD560). The kit included actin as internal control (IC), and samples with IC D values of <0.4 were classified as failed. The LAMP primer mix contained primers targeting the E and N gene for viral detection, and samples with D>0.4 were considered positive. Samples tested via qPCR (using 2 µl RNA as input) were processed using TaqMan Fast Virus 1-step Master mix, which uses the N1 primer set.

### Data processing

Demultiplexed FASTQ files were processed using umi_tools ^57^ (`umi_tools extract`) to only preserve reads with expected sample well barcodes (maximum 96). Reads were mapped to the three expected amplicon sequences with bwa ^58^ (`bwa mem`). Sorted BAM files were filtered to remove primer dimer reads and potential other noise. The number of UMIs as well as the number of reads per UMI was counted and merged for all the samples for downstream analysis. For downsampled data, we sampled a fraction of reads per plate at random with sam-tools ^59^ command `samtools view -s`. Using the EMBL cluster (run on Intel Xeon Gold 6136 CPU and 32 Gb of RAM), this pipeline allows generating count matrices in under 10 minutes for a sequencing run. Exact running time depends on available hardware and sequencing depth. For instance, while it takes about eight minutes for a single plate (FASTQ file) with 15 million reads to be processed, analysis time is reduced to 9 seconds when reads are down-sampled to 15,000. Downstream analysis of count tables was performed with the R programming language ^60^ using tidyverse ^61^ for data handling and ggplot2^62^ for visualization. Scripts for processing of sequencing data have been deposited at http://github.com/gtca/McQ.

## Data Availability

Software and Supplementary Tables are available at the shared github link. Supplementary Figures and Files are provided as part of the manuscript file.

http://github.com/gtca/McQ

## Acknowledgements

The construct encoding the MMLV_mut reverse transcriptase was a kind gift from Dr. Louise Walport and Dr. Svend Kjaer (Francis Crick Institute, London). We thank Alexander Aulehla for contributing to the overall design of the study, the management of early positive sample access and support in the implementation of routine testing at EMBL. D.B., M.F. and J. K. were supported by the EMBL International PhD program. D.B. was supported by a Darwin Trust fellowship. We are grateful to Alessandra Reversi and Sandra Clauder for project management to implement saliva testing at EMBL. We would like to thank the team of EMBL volunteers for pipetting samples for routine saliva testing at EMBL. We thank Simon Anders, the Genome Biology Computational Support team, Jan Provaznik, Jonathan Landry and Tobias Rausch for computational support for sample management. Research in the laboratory of A.R.K and L.S. was supported by core funding of the European Molecular Biology Laboratory. M.K. and H.G.K received funding through a research grant from the state of Baden-Württemberg (MWK).

## Author contributions

A.R.K., V.B., K.R., and A.T. conceived and coordinated the overall project. S.C.V conceived, designed and was responsible for coordination and implementation of the McQ NGS-based multiplexing approach. S.C.V. and J.K. performed experiments to establish and benchmark the McQ workflow. D.B. developed computational analysis pipelines to generate UMI count tables from McQ sequencing data, and D.B. and M.F. analysed data. S.C.V and A.R.K. advised analysis of McQ data. V.B. coordinated and performed sequencing of McQ data.V.B. and E.C. designed and coordinated implementation of automated RNA extraction. E.C., M.O, F.J. and J.Z. performed experiments to establish automated RNA extraction. K.R. coordinated production of homemade enzymes, and J.F. and N.D. expressed and purified homemade enzymes. N.D. advised optimization of reaction conditions with homemade enzymes and provided buffers. V.B. coordinated optimization of MMLV_mut reaction conditions. R.K., L.V. and K.M. designed and performed experiments to optimize reaction conditions for MMLV_mut reverse transcriptase. F.J. implemented automation of the McQ reverse transcription step on the Biomek i7 platform. L.V., F.J. and M.O. processed saliva samples for Figure 4. H.G.K collected and characterised RNA of clinical samples for establishing McQ. S.G. and A.H. collected and provided clinical swab samples for establishing bead-based RNA extraction. M.K. characterised samples used for benchmarking McQ. A.R.K., V.B. K.R. A.T. and L. M. S. advised the study. S.C.V., D.B. and A.R.K. wrote the manuscript. All authors read, edited and approved the final manuscript. The authors declare no conflict of interest.

## Supplementary Figures and Material for

**Figure S1.**
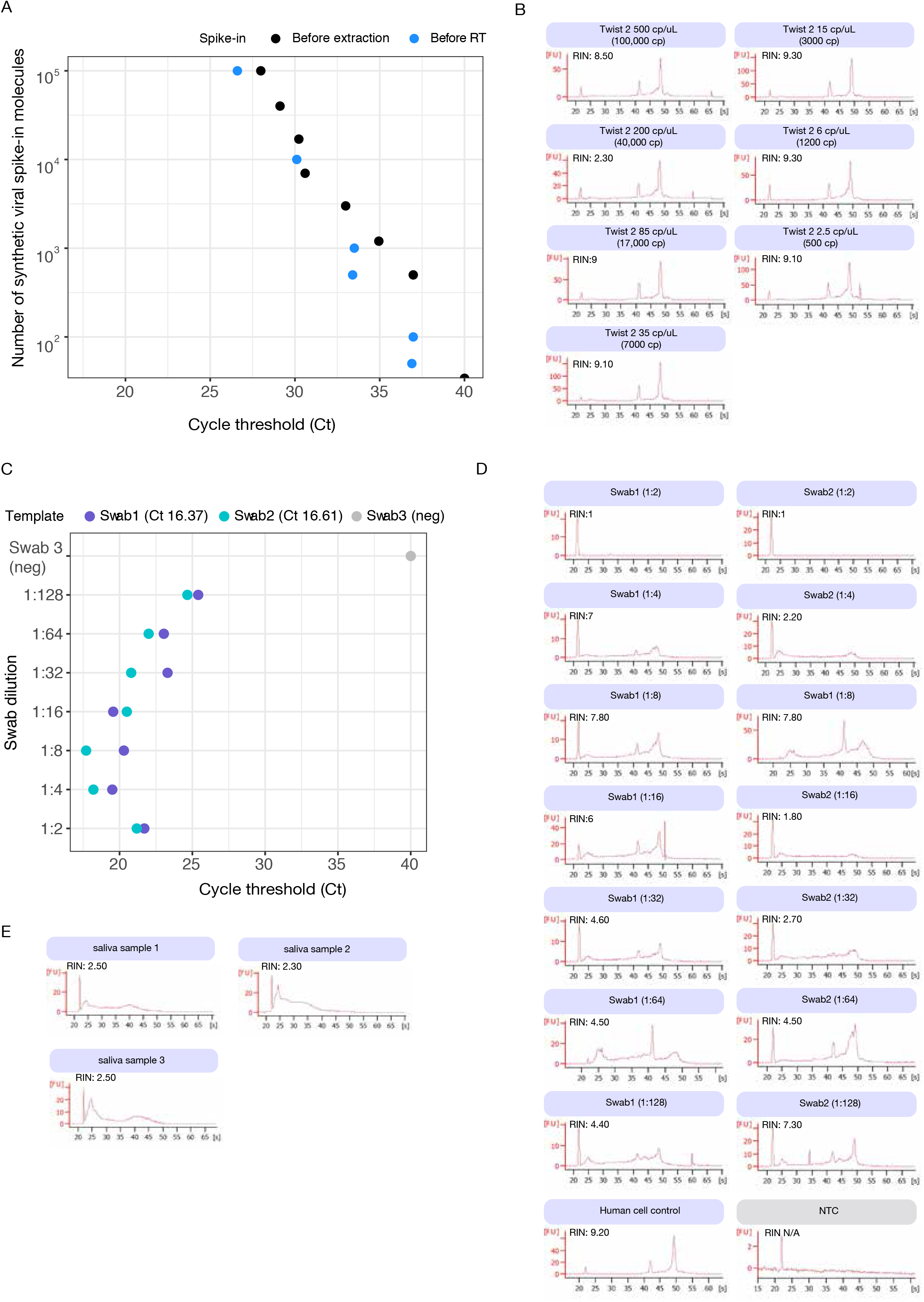
RNA extraction benchmarking. (A) Scatter plot depicts Ct values obtained in RT-qPCR for defined amounts of synthetic SARS-CoV-2 template added to samples before RNA extraction (spiked into Expi293F cell suspension at 106 cells/ml in saline solution) or directly added prior to RT. (B) Bioanalyzer traces and RNA integrity numbers (RIN) for extracted Expi293F cell suspension (200 µl) spiked with indicated amounts of synthetic SARS-CoV-2 template. (C) Scatter plot depicts Ct values obtained in RT-qPCR for dilutions of two swab samples extracted manually with our magnetic bead-based RNA extraction workflow that were previously tested positive via qPCR. (D) Bioanalyzer traces and RNA integrity numbers (RIN) for extracted swab samples diluted prior to extraction. NTC = no template control. Human cell control is a suspension of cells without viral RNA spiked in. (E) Bioanalyzer traces and RNA integrity numbers (RIN) for saliva samples extracted with our automated workflow.

**Figure S2.**
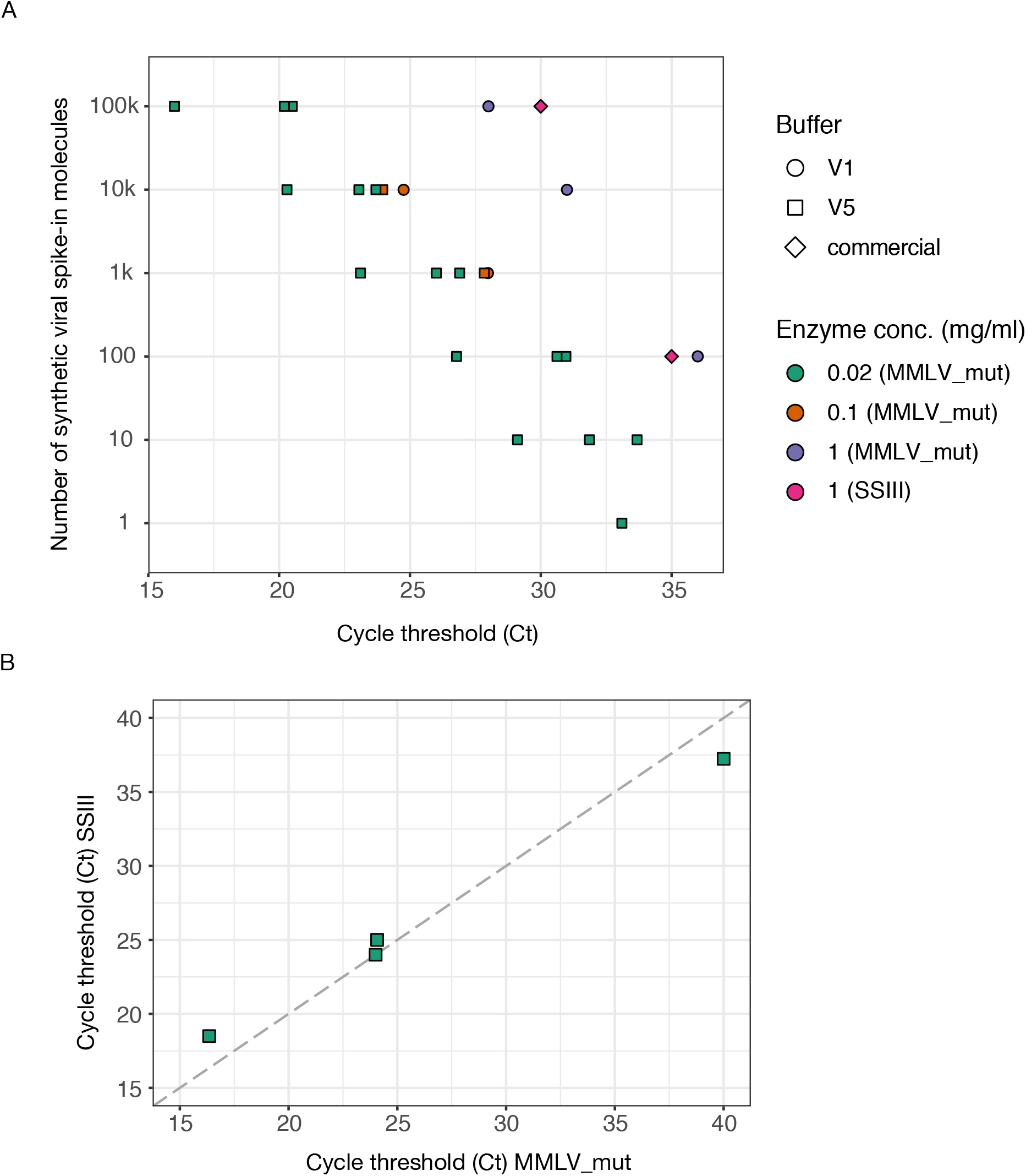
Establishing MMLV_mut reaction conditions. (A) Scatter plot of Ct values obtained for defined amounts of synthetic viral RNA template spiked into reactions. Colors label different MMLV_mut enzyme concentrations used in reverse transcription (stocks of 0.02 mg/ml, 0.1 mg/ml or 1 mg/ml) and shapes encode different buffers. (B) Comparison of Ct values obtained for four clinical samples with MMLV_mut using optimized conditions (buffer V5 and 0.02 mg/ml enzyme stock) and SYBR Green Ready mix, and Ct values obtained from RT-qPCR assays run in a clinical diagnostics facility using a commercially available test kit.

**Figure S3.**
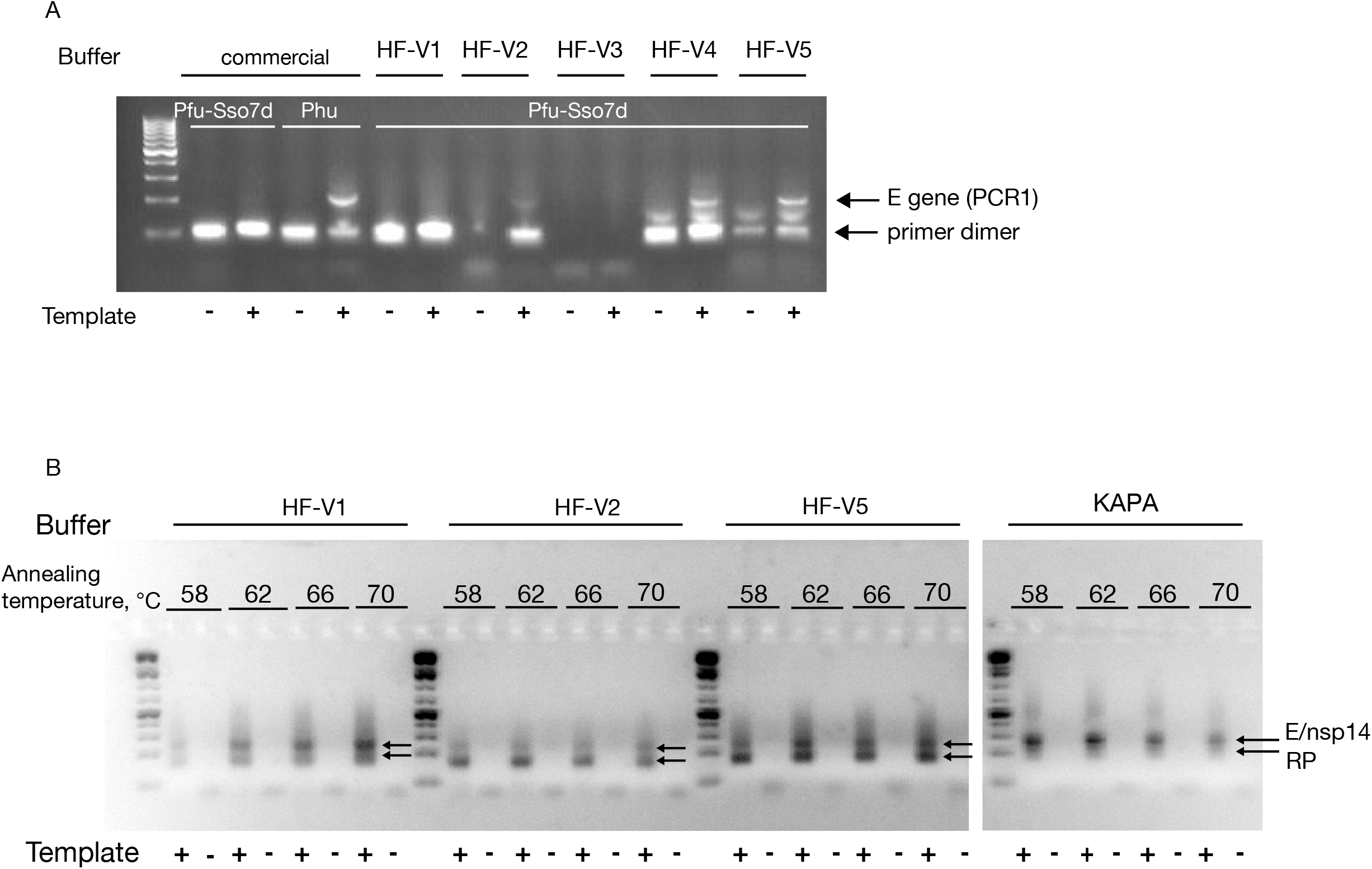
Performance of Pfu-Sso7d polymerase in different reaction buffers. (A) Gel shows E amplicon after PCR1 (216 bp) amplified with homemade Pfu-Sso7d in different reaction buffers (HF V1-V5) and with commercial Phusion polymerase (Phu). PCRs were performed on half of the cDNA reverse transcribed from 10,000 Twist synthetic RNA templates (using E-specific RT primer, RT buffer V1 and 1 mg/ml MMLV_mut) (+). Non-template controls (-) contain nuclease-free water instead of cDNA. Arrows indicate E amplicon and primer dimer bands. (B) Buffer comparison and annealing gradient for PCR2 with Pfu-Sso7d and commercial KAPA polymerase. PCRs were performed on PCR1 products of mixtures of 10,000 synthetic viral RNA molecules and 100 ng human total RNA reverse transcribed with a mixture of E, nsp14 and RP RT primers (using RT buffer V5 + Tre and 0.1 mg/ml MMLV_mut) (+). Non-template controls (-) received nuclease-free water instead of cDNA.

**Figure S4.**
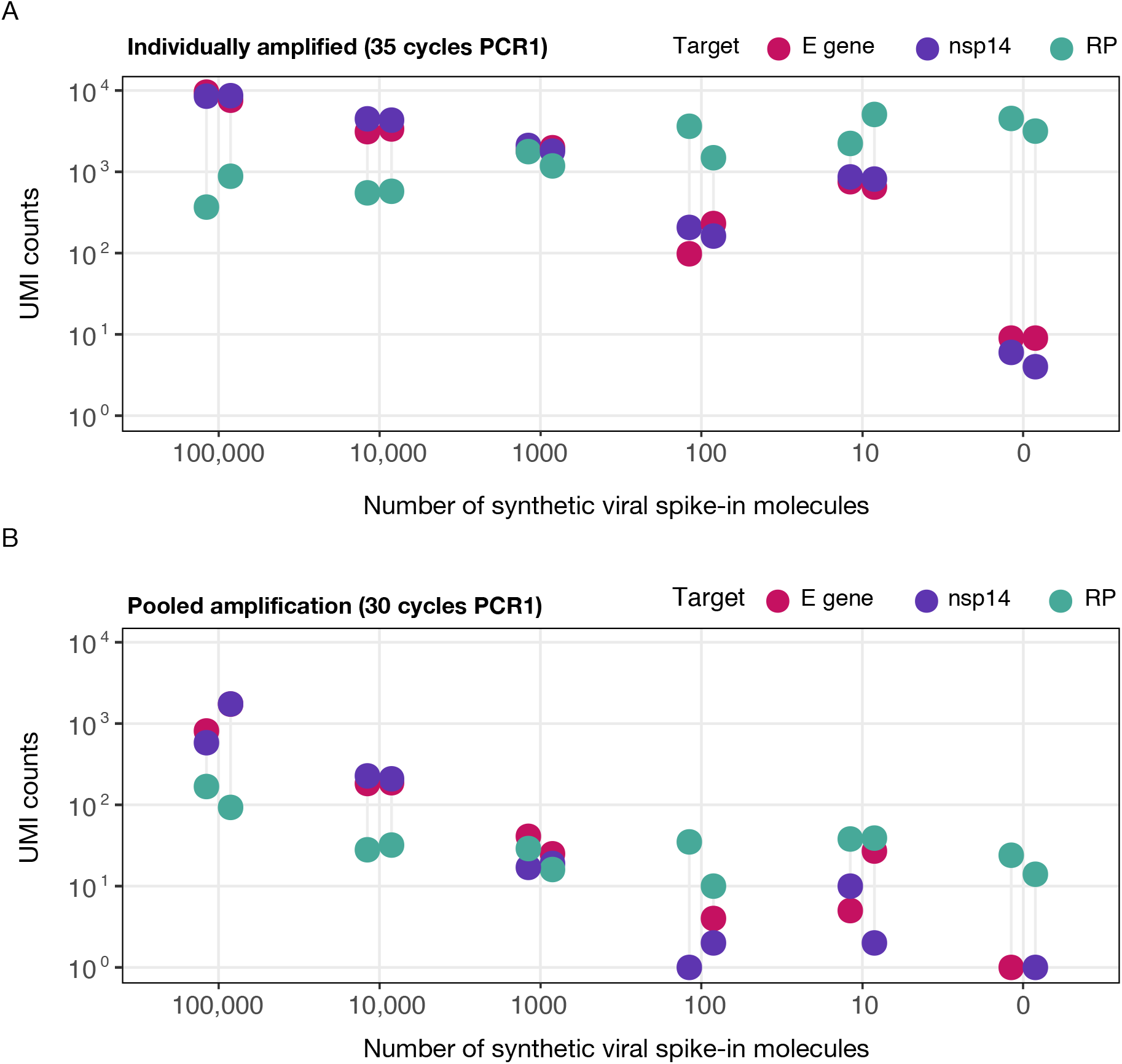
McQ quantitatively detects synthetic viral RNA templates. Twelve samples containing defined numbers of synthetic SARS-CoV-2 RNA template (0-100,000 copies) and human RNA were reverse transcribed (using buffer V1 and 1 mg/ml MMLV_mut stock) and amplified individually (A) or in a pool of twelve (B) using the McQ workflow. Scatter plots depict the number of Unique Molecular Identifiers (UMI) labeling viral targets (E gene: pink, nsp14: purple) or human control (RP: green) as a function of the number of synthetic viral RNA molecules spiked into 10 ng total human RNA. Each viral spike-in amount was processed in duplicate. UMI counts reflect unique RNA templates present in the sample.

**Figure S5.**
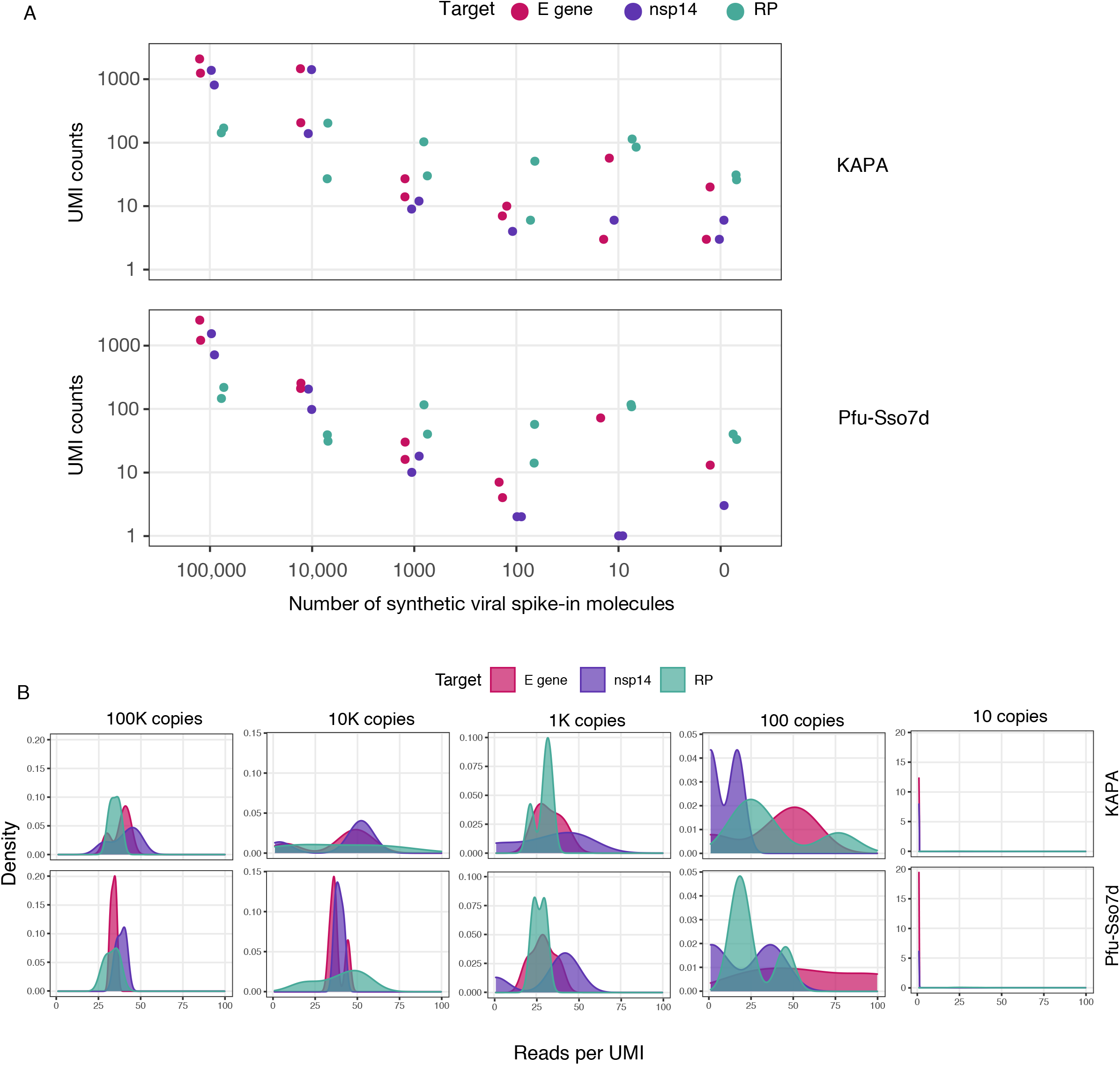
Benchmarking of homemade Pfu-Sso7d polymerase against commercial KAPA polymerase in PCR2. (A) Scatter plots show the number of UMI detected for viral targets (E gene: pink, nsp14: purple) or human control (RP: green) for mixtures of 10 ng human total RNA with indicated amounts of viral template with commercial KAPA (upper panel) or homemade Pfu-Sso7d (lower panel) polymerase in PCR2. Pfu-Sso7d was used in PCR1 for both. Each viral spike-in amount was processed in duplicate. (B) Density plots depict distributions of read numbers per UMI for samples containing mixtures of 10 ng human total RNA with indicated numbers of synthetic viral templates. Samples containing defined numbers of synthetic SARS-CoV-2 RNA template (0-100,000 copies) and 10 ng human RNA were processed in pools of twelve using the McQ workflow and using either commercial KAPA (upper panels) or Pfu-Sso7d (lower panels) polymerase in PCR2. Density plots are drawn separately for different targets (E gene: pink, nsp14: purple or human control (RP): green).

**Figure S6.**
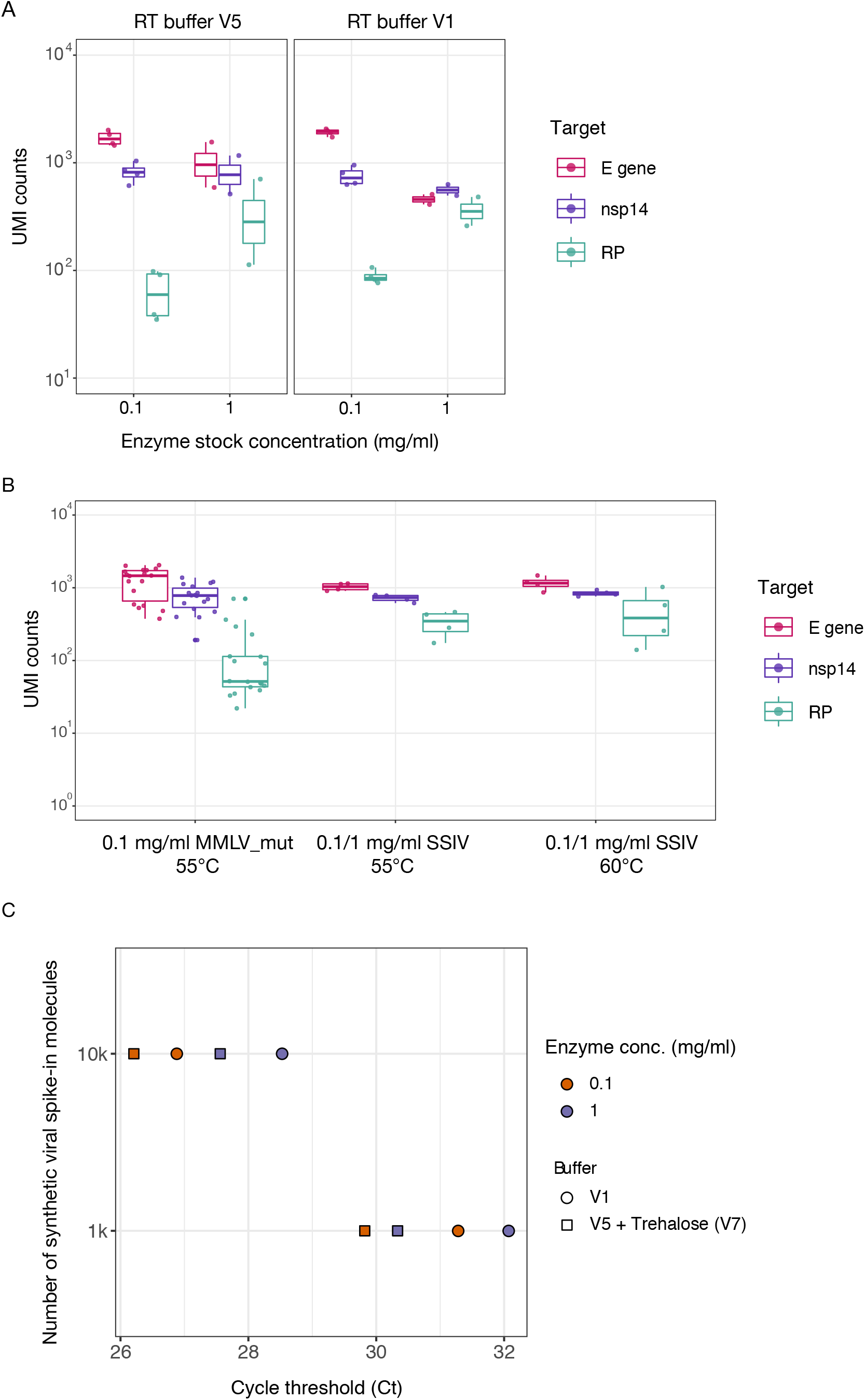
Effect of different reverse transcription conditions and enzymes on UMI recovery. (A) Boxplots depict the number of UMI detected for viral targets (E gene: pink, nsp14: purple) or human controls (RP: green) with different reverse transcription buffers and MMLV_mut enzyme concentrations. 10,000 Twist synthetic SARS-CoV-2 RNA templates and 10 ng human RNA was used as input into reverse transcription for each sample. (B) Same as A but comparing MMLV_mut to commercial SSIV reverse transcriptase at two concentrations (using 1 µl and 0.1 µl enzyme) and RT temperatures. (C) Scatter plot of Ct values obtained for defined amounts of synthetic viral RNA template spiked into RT reactions (using the McQ E primer set). qPCR was performed using SYBR Green Ready Mix. Colors label different MMLV_mut enzyme concentrations used in reverse transcription (stocks of 0.1 mg/ml or 1 mg/ml) and shapes encode different buffers.

**Figure S7.**
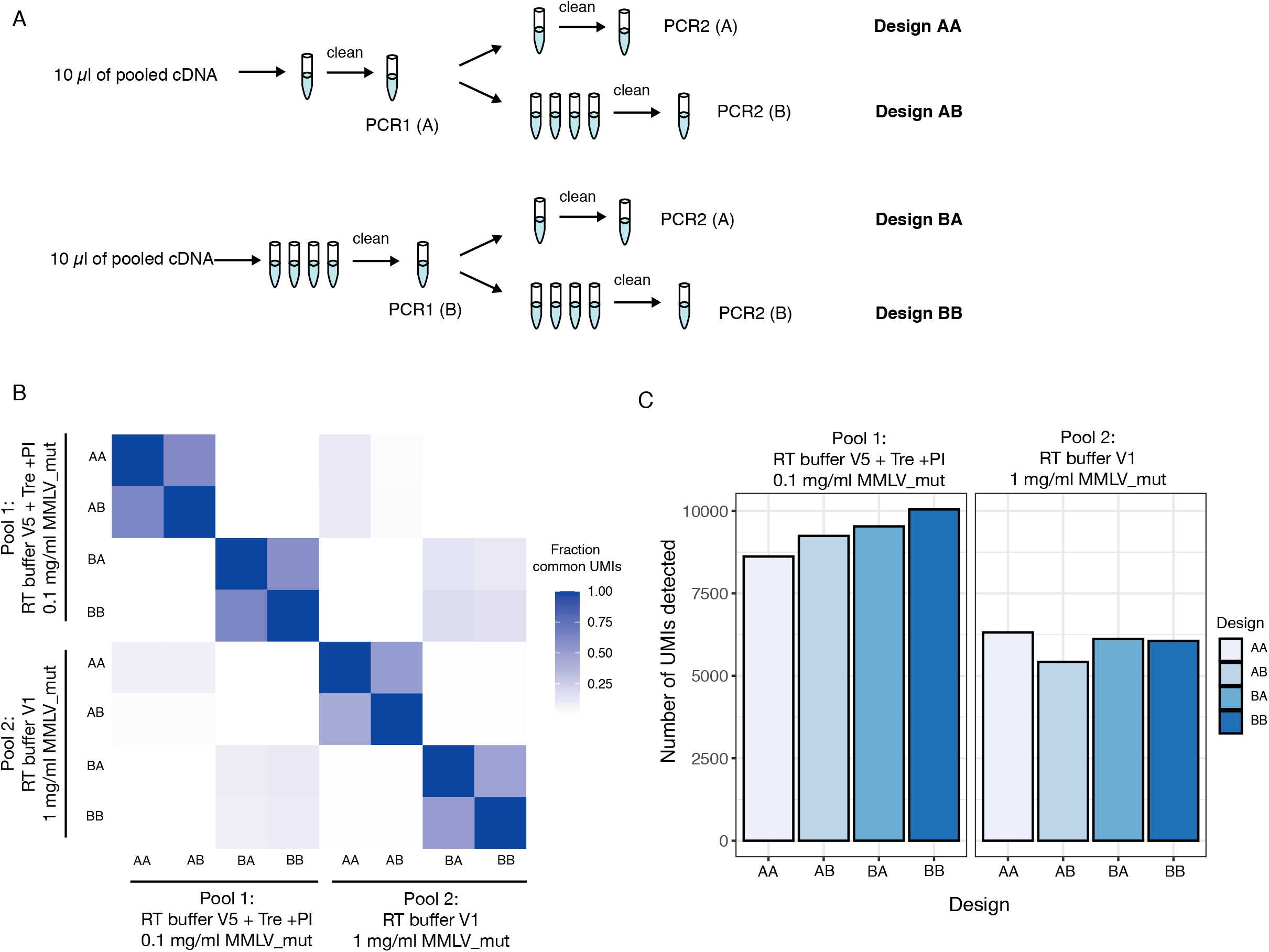
PCR jackpotting reduces UMI recovery and diversity. (A) Twelve samples containing defined numbers of synthetic SARS-CoV-2 RNA template (0-100,000 copies) and 10 ng human RNA were reverse transcribed and pooled for PCR amplification. Four different amplification designs were tested to identify PCR-related factors limiting UMI recovery. Samples AA and AB result from the same PCR1 as do samples BA and BB. (B) Heatmap of correlation between samples based on detected UMIs. Color indicates the fraction of UMIs that are shared between two samples. Samples were reverse transcribed in indicated conditions prior to PCR amplification. (C) Number of UMI recovered with each design (representing different PCR amplification strategies) for each pool (representing different reverse transcription conditions).

**Figure S8.**
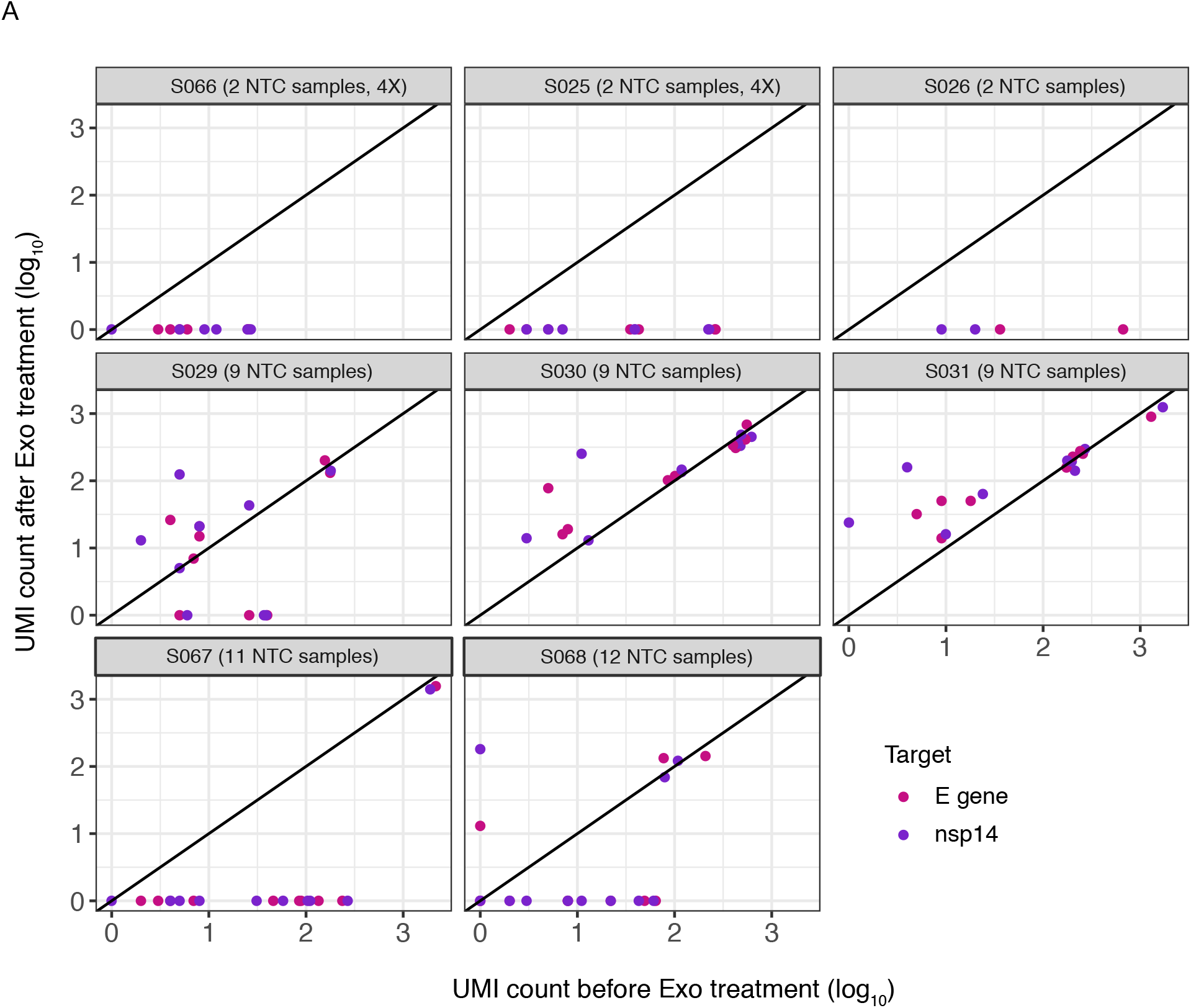
Free-floating index primers are a source of background reads. Scatterplots depict number of UMI detected for viral E (pink) and nsp14 (purple) targets in non-template control (NTC) samples (containing only human RNA i.e. representing background levels) in the original samples and the same samples sequenced after an additional Exonuclease I treatment after PCR2 to remove excess index primers. Black line indicates a slope of 1. Sample pool ID and number of NTC samples present in each sample pool are indicated in grey boxes above panels. Sample pools S025 and S066 were present in four variations, but each with identical sample composition.

**Figure S9.**
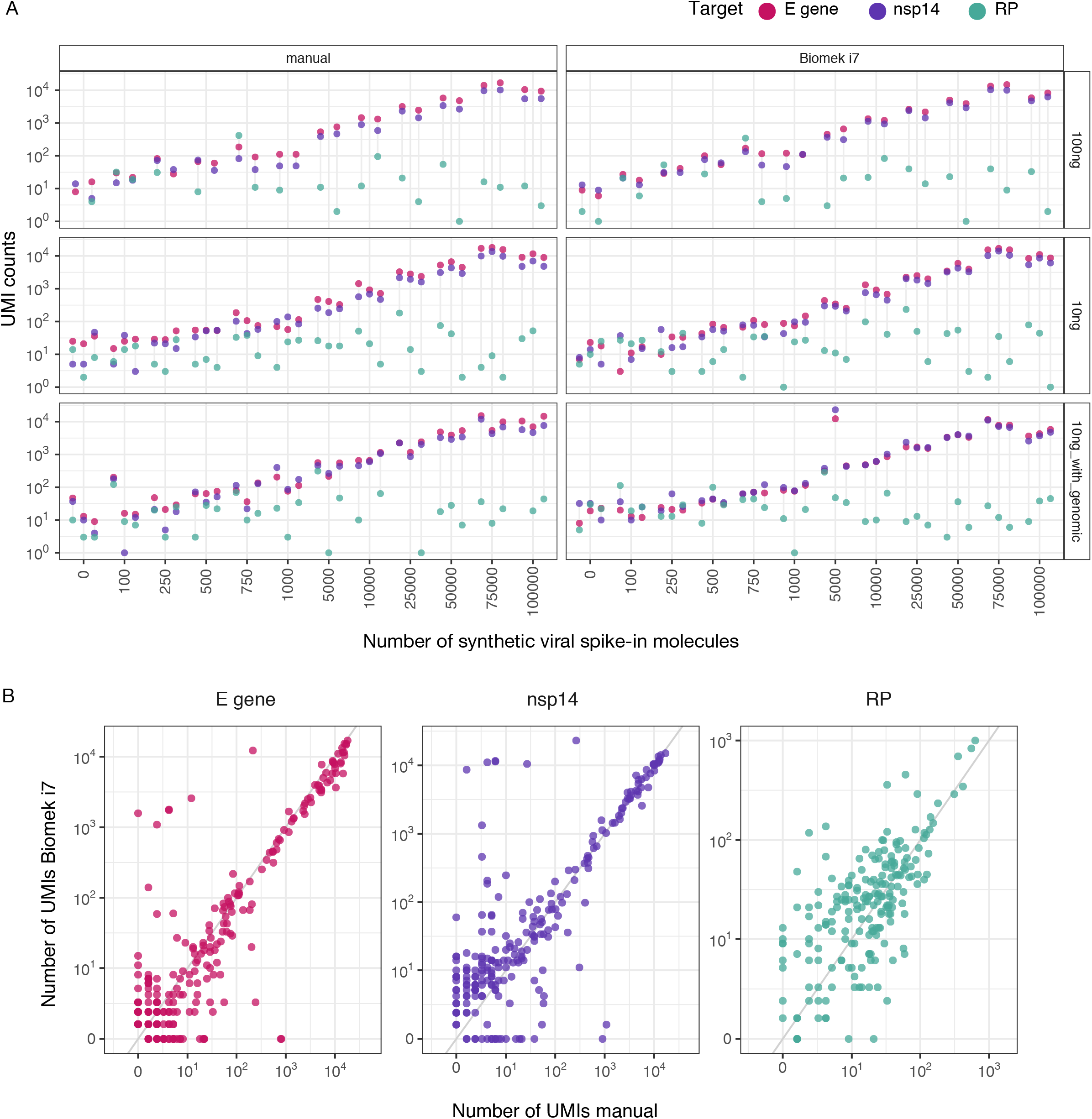
Benchmarking of McQ RT step on an automated Biomek i7 platform. (A) 96 samples containing defined numbers of synthetic SARS-CoV-2 RNA template (0-100,000 copies) and human RNA were processed using the McQ workflow and performing RT manually (left, using multichannel pipettes) or on an automated Biomek i7 platform (right, protocol described in File S3). Scatter plots depict the number of UMI labeling viral targets (E gene: pink, nsp14: purple) or human control (RP: green) as a function of the number of synthetic viral RNA molecules spiked into indicated amounts of total human RNA. Each viral spike-in amount was processed with varying amounts of human RNA mixed into the sample (100 ng RNA, 10 ng RNA, 10 ng RNA + 10 ng genomic DNA) in duplicate or triplicate. UMI counts reflect unique RNA templates present in the sample. (B) Comparison of number of UMI detected for each of the 96 samples processed manually or on the i7 platform.

**Figure S10.**
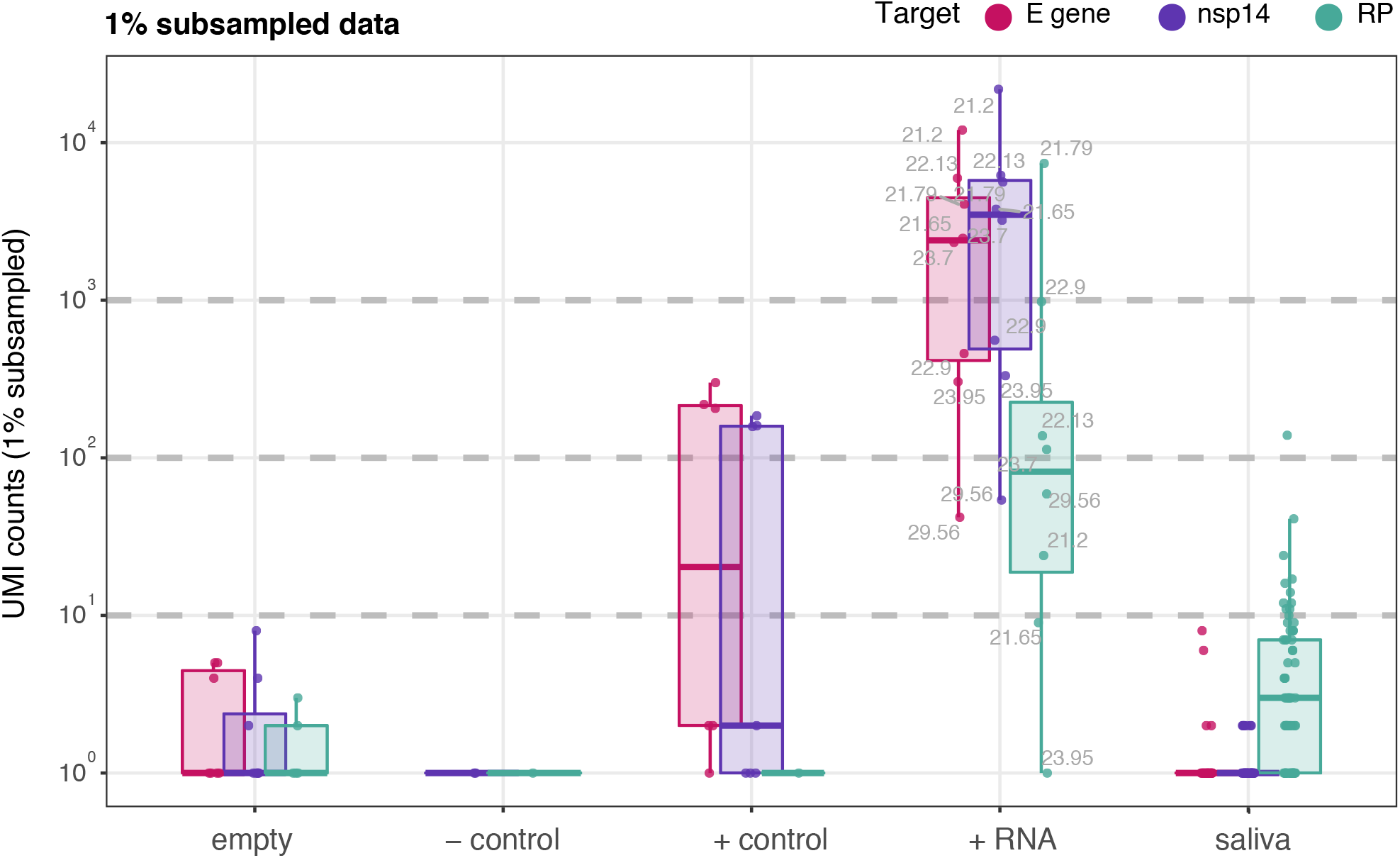
Detection of SARS-CoV-2 RNA in extracted saliva samples (1% subsampled data). Scatter plots show the number of UMI detected for viral targets (E gene: pink, nsp14: purple) or human control (RP: green) in RNA extracted from 136 saliva samples provided by healthy volunteers (saliva). Empty = water, added before RNA extraction, negative control = saliva from a previously tested negative sample, positive control = highly diluted negative saliva sample spiked with RNA from a sample tested positive for SARS-CoV-2 and100,000 molecules of Twist synthetic RNA (in water, spiked in prior to extraction), positive RNA = extracted RNA from samples tested positive for SARS-CoV-2 (added after RNA extraction). Boxplots are overlaid to visualize distributions. Data is the same as Figure 4 except only 1% of the data (corresponding to approximately 2600 reads/sample) is plotted.

### Supplementary File 1: Automation of RNA extraction on the Biomek i7 liquid-handling platfor

The protocol follows the <u>original protocol</u> by BOMB.Bio^1^ adapted for an automated implementation on the Biomek i7. This protocol can in principle be implemented on any liquid handling robot equipped to work with magnetic beads. The input of the automated procedure is a heat-inactivated sample in GITC lysis buffer, manually formatted in 96 deep-well plates from Abgene. Individual steps as programmed on the Biomek i7 are described in Figure 1 and deck layout at start is shown in Figure 2. Table 1 lists required consumables.

**Figure 1.**
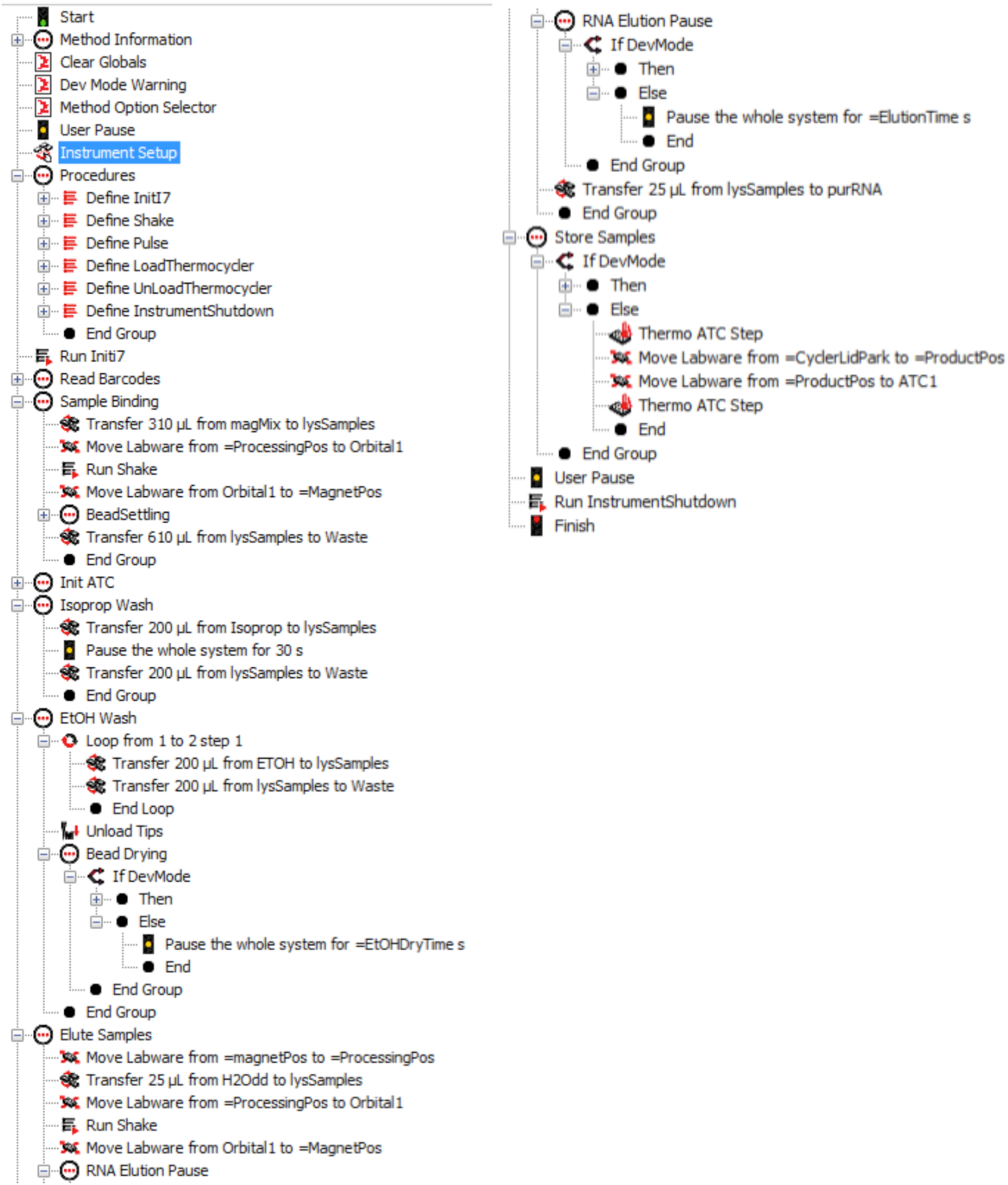
Workflow for automated RNA extraction implemented on the i7 platform. The figure shows a detailed step-by-step workflow of the protocol used for extraction on the i7. We start the extraction protocol with a plate containing 300ul (sample + lysis-buffer). The sample in the end is then eluted in 25ul into a PCR-plate.

**Figure 2.**
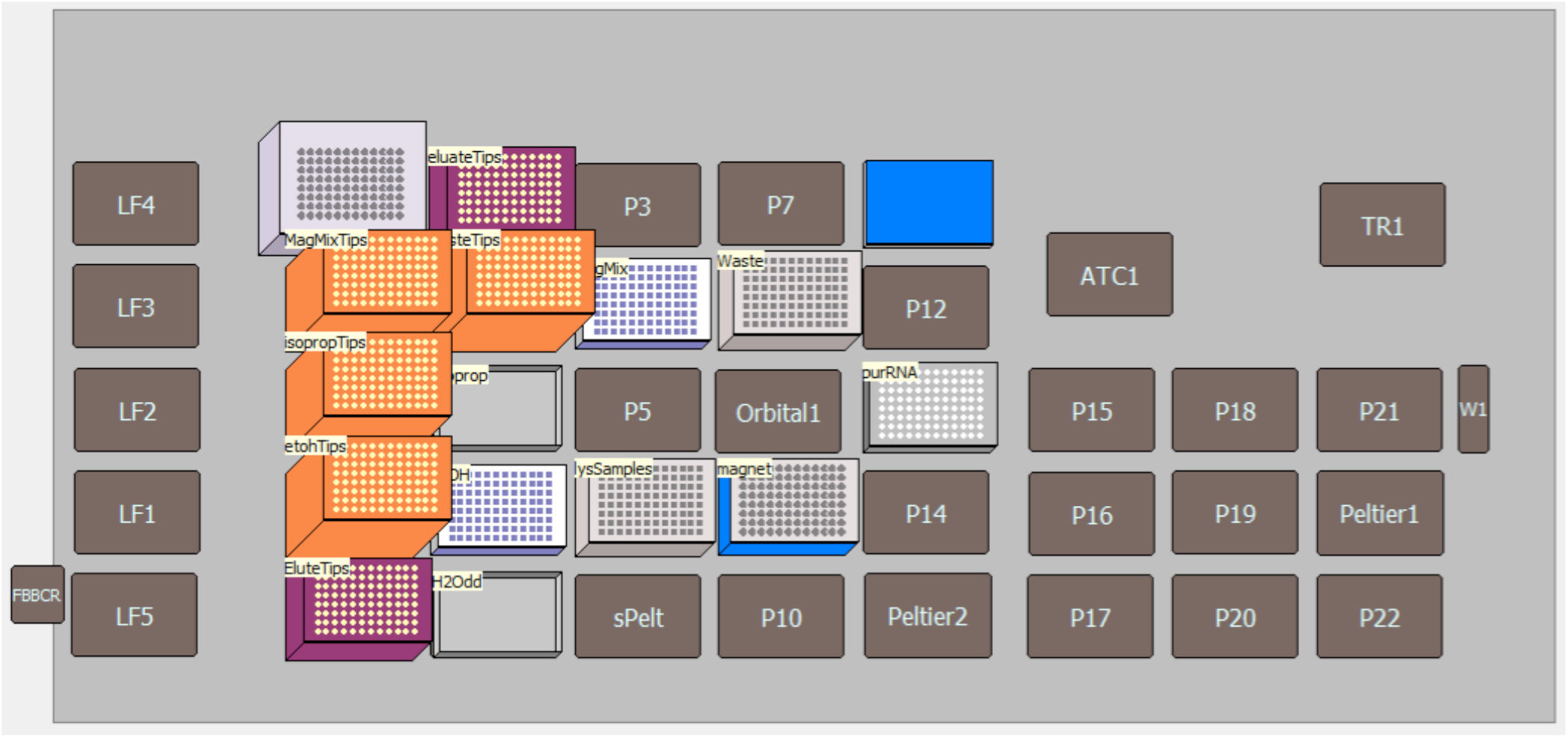
Deck-layout of the i7 system for RNA extraction. Pink boxes are 50ul Beckman tip-boxes, orange boxes are 1000 ul Beckman tip-boxes. “Isoprop” and “H20dd” are upside-down tip-box lids used as reservoirs. Waste and “lysSamples” are AB-0932 plates. “MagMix” and “EtOH” are AB-1127 plates. The Alpaqua Magnum-EX is used as magnet. Samples are eluted in “purRNA”, which is a barcoded PCR-plate (Thermo). The blue position is a metal PCR-lid used for storing the samples in the cycler after extraction.

**Table 1:** Consumables.

| *Material* | *Provider* | *Order #* | *Number used* |
| --- | --- | --- | --- |
| 50 ul tips | Beckman | B85888 | 2 |
| 1000 ul tips | Beckman | B85955 | 5 |
| PCR-plates | ThermoFischer |  | 1 |
| Tip-box lids | Beckman | - | 2 |
| AB-1127 | Axon | 26032 | 2 |
| AB-0932 | ThermoFischer | AB0932 | 2 |
| Magnet Magnum EX | Alpaqua | - | 1 |
| PCR-Lid | - | - | 1 |

### GITC buffer

6 M GITC

50 mM Tris-HCl pH 7.6-8

2% Sarcosyl

20 mM EDTA

0.1% Octanal

Silica coated beads (commercial): Serasil-Mag™ 400 beads (# 29357371; Cytiva - Formerly GE Healthcare Life Sciences). Note: 400 means diameter is 400nM._Beads were prepared following the instructions in the BOMB.Bio protocol. The rest of the reagents needed for this protocol are ethanol (70%) and isopropanol.

### Supplementary File 2: Expression and purification of homemade enzymes

#### Expression and purification of MMLV_mut reverse transcriptase

The construct encoding the MMLV_mut reverse transcriptase was a kind gift from Dr. Louise Walport and Dr. Svend Kjaer (Francis Crick Institute, London). The pET28a MMLV_mut plasmid was transformed freshly into *E. coli* BL21(DE3) cells (Stratagene). Cells were grown in TB-FB auto-induction medium (TB-FB + 0.05% glucose + 1.5% lactose + 2 mM MgSO_4_) supplemented with 30 µg/ml kanamycin at 37°C until OD_600_ ∼ 0.9. The temperature was then lowered to 18°C and the cells were grown further overnight at 18°C. The next morning, the cells were harvested by centrifugation. The cell pellet was resuspended in lysis buffer (50 mM Tris-HCl pH 8.0 (4°C), 1 M NaCl, 20 mM imidazole, 10% glycerol, 1 mM PMSF and cOmplete EDTA-free protease inhibitors (Roche)) and lysed by 5 passages through an M-110L Microfluidizer processor (Microfluidics). After centrifugation (30 min 50,000 x g, 4°C), the cleared lysate was loaded onto a 5 ml HisTrap HP column (GE Healthcare) pre-equilibrated with running buffer (50 mM Tris-HCl pH 8.0 (4°C), 1 M NaCl, 20 mM imidazole, 10% glycerol). After washing the column with running buffer, the His_6_-tagged MMLV_mut reverse transcriptase was eluted with 25 mM Tris-HCl pH 8.0, 500 mM NaCl, 200 mM imidazole and 10% glycerol. To remove the affinity tag, thrombin protease (Merck) was added to the His_6_-MMLV_mut protein and the sample was incubated overnight at 8°C in cleavage buffer (20 mM Tris pH 8.0, 250 mM NaCl, 100 mM imidazole, 2.5 mM CaCl_2_, 10% glycerol). After thrombin cleavage, the sample was diluted 1:1 with 20 mM Tris pH 8.0, 10% glycerol to lower the amount of salt and facilitate binding to the ion exchange column. The diluted sample (20 mM Tris pH 8.0 (4°C), 125 mM NaCl, 50 mM imidazole, 1.25 mM CaCl_2_, 10% glycerol) was loaded onto a 5 ml HiTrap Q HP and a 5 ml HiTrap SP HP (GE Healthcare) column that were coupled in tandem. After washing both columns with 20 mM Hepes pH 7.0, 100 mM NaCl and 10% glycerol, the HiTrap Q HP column was removed and the HiTrap SP HP column was eluted in a gradient to 20 mM Hepes pH 7.0, 1 M NaCl and 10% glycerol over 12 column volumes. Finally, the MMLV_mut protein was loaded onto a HiLoad 16/600 Superdex 200 pg (GE Healthcare) size exclusion chromatography column equilibrated with SEC buffer (40 mM Tris-HCl pH 7.5 (25°C), 200 mM KCl). The fractions containing the MMLV_mut enzyme were pooled and concentrated to 4.3 mg/ml. The final protein was stored at -20°C in 20 mM Tris Ph 7.5 (25°C), 100 mM KCl, 1 mM DTT, 0.1 mM EDTA, 0.01% NP40 and 50% glycerol at a concentration of 2 mg/ml. The yield obtained from 2 liters of expression culture amounted to 120 mg of MMLV_mut reverse transcriptase.

#### Expression and purification of Pfu-Sso7d polymerase

The pET-His_10_-GS-TEV-Pfu-Sso7d construct was transformed into *E. coli* Rosetta™ 2 (DE3) (Novagen) cells. Cells were grown in TB-FB auto-induction medium supplemented with 30 µg/ml kanamycin and 34 µg/ml chloramphenicol at 37°C until OD_600_ ∼ 0.8. The temperature was then lowered to 18°C and the cells were grown further overnight at 18°C. The next morning, the cells were harvested by centrifugation. The cell pellet was resuspended in running buffer (50 mM Tris-HCl pH 8.0 (4°C), 1 M NaCl, 20 mM imidazole, 10% glycerol, 1 mM PMSF (Sigma) and cOmplete EDTA-free protease inhibitors (Roche)) and lysed by 5 passages through a microfluidizer device. Next, the lysate was heat-shocked for 45 minutes at 80°C before being cooled down on ice. After centrifugation (30 min., 50000 x *g*, 4°C), the cleared lysate was loaded onto two 1 ml HisTrap HP columns (GE Healthcare) that were coupled in tandem and pre-equilibrated with running buffer (50 mM Tris-HCl pH 8.0 (4°C), 1 M NaCl, 20 mM imidazole and 10% glycerol). After washing with running buffer, the His_10_-tagged Pfu-Sso7d polymerase was eluted with 50 mM Tris-HCl pH 8.0 (4°C), 150 mM NaCl, 400 mM imidazole and 10% glycerol. To remove the affinity tag, His_10_-tagged TEV protease was added to the sample in a 1:100 ratio and the mixture was then diluted 10-fold with 20 mM Tris pH 8.0 (4°C), 100 mM NaCl and 10% glycerol. The cleavage reaction was allowed to continue for 4 hours at room temperature. After TEV cleavage, the sample was loaded onto a 5 ml HiTrap Heparin HP column (GE Healthcare) pre-equilibrated with 20 mM Tris pH 8.0 (4°C), 100 mM NaCl and 10% glycerol. After washing, the Heparin column was eluted in a gradient to 20 mM Tris pH 8.0 (4°C), 500 mM NaCl and 10% glycerol over 40 column volumes. Finally, the untagged Pfu-Sso7d polymerase was loaded onto a HiLoad 16/600 Superdex 200 pg (GE Healthcare) size exclusion chromatography column equilibrated with SEC buffer (20 mM Tris-HCl pH 7.5 (25°C), 100 mM KCl). The fractions containing the Pfu-Sso7d polymerase were pooled and concentrated to 2.2 mg/ml. The final samples were stored at -20°C in 20 mM Tris pH 7.5 (25°C), 100 mM KCl, 1 mM DTT, 0.1 mM EDTA, 200 µg/ml BSA and 50% glycerol at a concentration of 1 mg/ml. The yield obtained from 1 liter of expression culture amounted to 17 mg of Pfu-Sso7d polymerase. Titration of Pfu-Sso7d against commercial Phusion polymerase revealed that 0.5 µl (0.07 mg/ml) Pfu-Sso7d per 50 µl reaction performed the same as 1U commercial Phusion.

#### MMLV_mut storage Buffer

20 mM Tris-HCl, pH 7.5 @ 25°C

100 mM KCl

1 mM DTT

0.1 mM EDTA 0.01% NP-40 50% Glycerol

#### MMLV_mut reaction buffers

**MMLV\_mut reaction buffers:**
| 1x MMLV_mut buffer | 5 x MMLV_mut buffer |
| --- | --- |
| 50 mM Tris-HCl,<br>pH 8.3 @ 25°C<br>75 mM KCl<br>6 mM MgCl<sub>2</sub> | 250 mM Tris-HCl,<br>pH 8.3 @ 25°C<br>375 mM KCl<br>30 mM MgCl<sub>2</sub> |

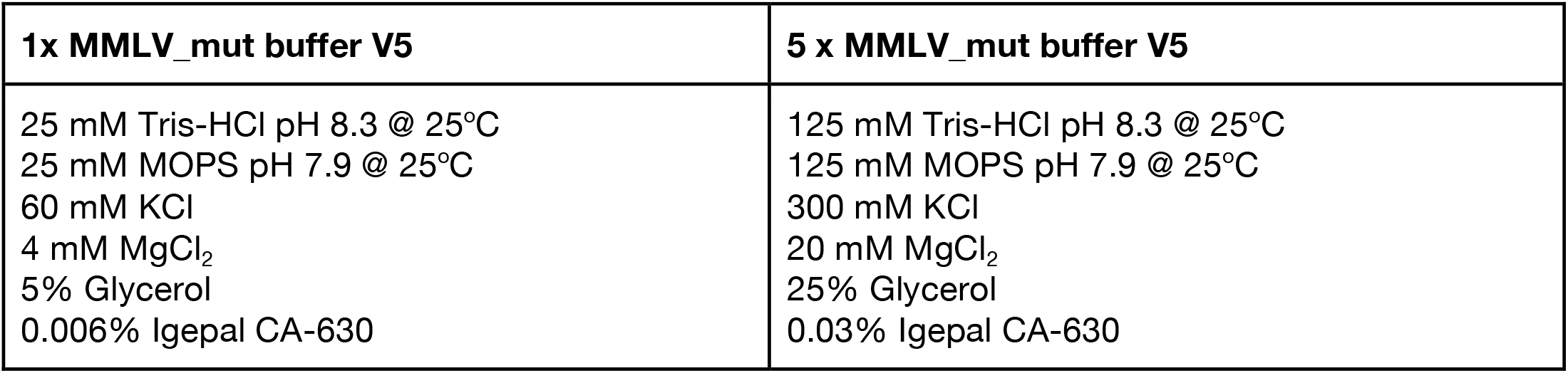

| **1x MMLV_mut buffer V7** | **5 x MMLV_mut buffer V7** |
| --- | --- |
| 25 mM Tris-HCl pH 8.3 @ 25°C<br>25 mM MOPS pH 7.9 @ 25°C<br>60 mM KCl<br>4 mM MgCl<sub>2</sub><br>200 mM Trehalose<br>5% Glycerol<br>0.006% Igepal CA-630 | 125 mM Tris-HCl pH 8.3 @ 25°C<br>125 mM MOPS pH 7.9 @ 25°C<br>300 mM KCl<br>20 mM MgCl<sub>2</sub><br>1000 mM Trehalose<br>25% Glycerol<br>0.03% Igepal CA-630 |

#### Pfu-Sso7d storage Buffer

20 mM Tris-HCl, pH 7.5 @ 25°C

100 mM KCl

1 mM DTT

0.1 mM EDTA

200 μg/ml BSA

50% Glycerol

#### Pfu-Sso7d reaction Buffers

*Source: PepCore EMBL*

| **1x HF buffer V1** | **10x HF buffer V1** |
| --- | --- |
| 20 mM Tris-HCl pH 8.8 @ 25°C<br>60 mM KCl<br>10 mM (NH<sub>4</sub>)<sub>2</sub>SO<sub>4</sub><br>2 mM MgCl<sub>2</sub><br>0.1% Triton-100<br>0.01 mM EDTA | 200 mM Tris-HCl pH 8.8 @ 25°C<br>600 mM KCl<br>100 mM (NH<sub>4</sub>)<sub>2</sub>SO<sub>4</sub><br>20 mM MgCl<sub>2</sub><br>1% Triton-100<br>0.1 mM EDTA |

| **1x HF buffer V2** | **5x HF buffer V2** |
| --- | --- |
| 30 mM Tris-HCl (pH 10 @ 25°C)<br>10 mM KCl<br>10 mM (NH<sub>4</sub>)<sub>2</sub>OAc<br>2 mM MgSO<sub>4</sub><br>0.1% Triton X-100<br>0.1 mg/ml BSA | 150 mM Tris-HCl (pH 10 @ 25°C)<br>50 mM KCl<br>50 mM (NH<sub>4</sub>)<sub>2</sub>OAc<br>10 mM MgSO<sub>4</sub><br>0.5% Triton X-100<br>0.5 mg/ml BSA |

| **1x HF buffer V3** | **5x HF buffer V3** |
| --- | --- |
| 30 mM Tris-HCl (pH 10 @ 25°C)<br>40 mM K<sub>2</sub>SO<sub>4</sub><br>1 mM (NH<sub>4</sub>)<sub>2</sub>SO<sub>4</sub><br>2 mM MgSO<sub>4</sub><br>0.1% Triton X-100 | 150 mM Tris-HCl (pH 10 @ 25°C)<br>200 mM K<sub>2</sub>SO<sub>4</sub><br>5 mM (NH<sub>4</sub>)<sub>2</sub>SO<sub>4</sub><br>10 mM MgSO<sub>4</sub><br>0.5% Triton X-100 |

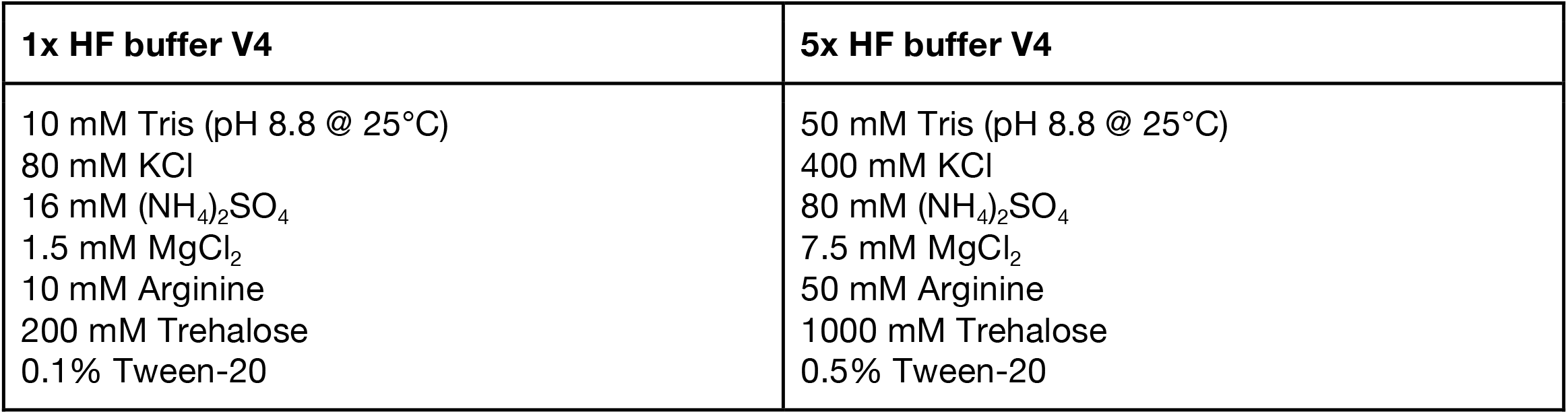

| **1x HF buffer V5** | **5x HF buffer V5** |
| --- | --- |
| 10 mM POPSO (pH 8.8 @ 25°C)<br>80 mM KCl<br>16 mM (NH<sub>4</sub>)<sub>2</sub>SO<sub>4</sub><br>1.5 mM MgCl<sub>2</sub><br>10 mM Arginine<br>200 mM Trehalose<br>100 mM Sarcosine<br>0.1% Tween-20 | 50 mM POPSO (pH 8.8 @ 25°C)<br>400 mM KCl<br>80 mM (NH<sub>4</sub>)<sub>2</sub>SO<sub>4</sub><br>7.5 mM MgCl<sub>2</sub><br>50 mM Arginine<br>1000 mM Trehalose<br>500 mM Sarcosine<br>0.5% Tween-20 |

Note: *Pfu-Sso7d* Buffers HF V2-V5 are from the following webpage: https://pipettejockey.com/2017/08/18/purifying-commonly-used-enzymes-homebrew-phusion/

**Figure 1.**
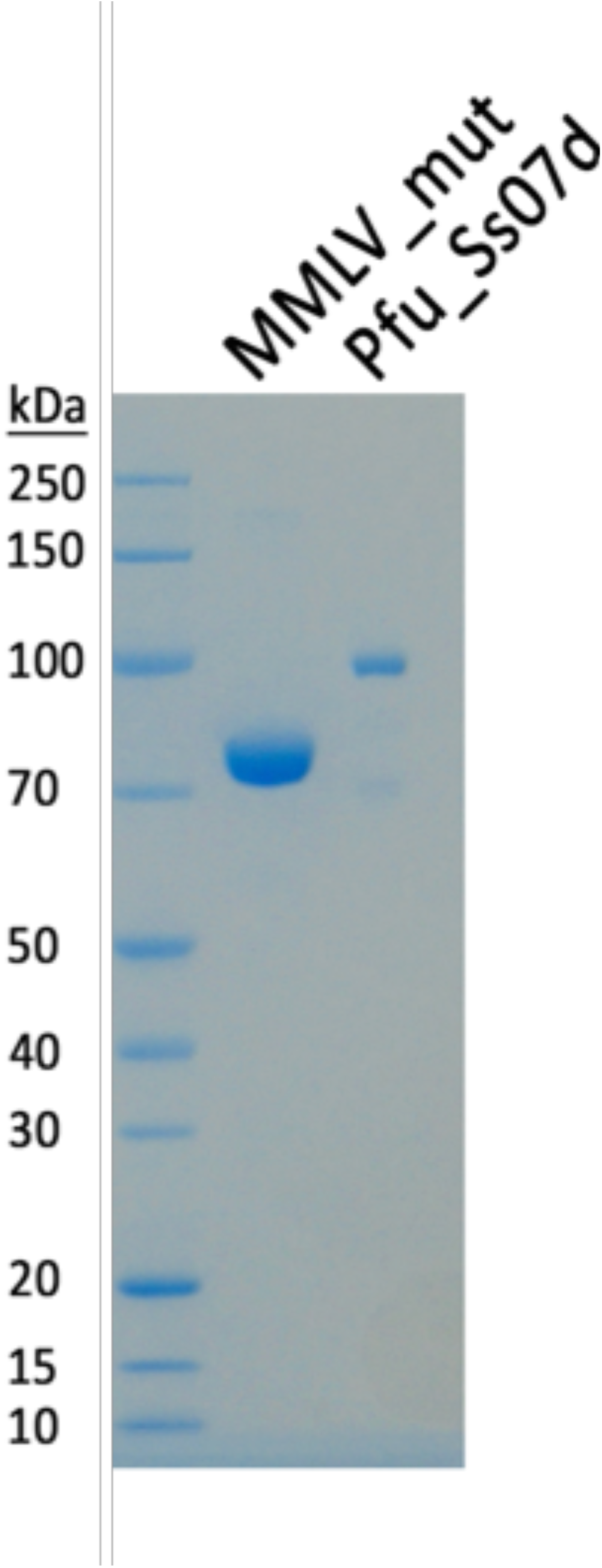
Purification of homemade enzymes. SDS-PAGE analysis of His_6_ purified MMLV_mut reverse transcriptase and Pfu-Sso7d polymerase after size exclusion chromatography. The single bands run at molecular weights of 76 kDA (MMLV_mut) and 98 kDa (Pfu-Sso7d).

**Figure 2.**
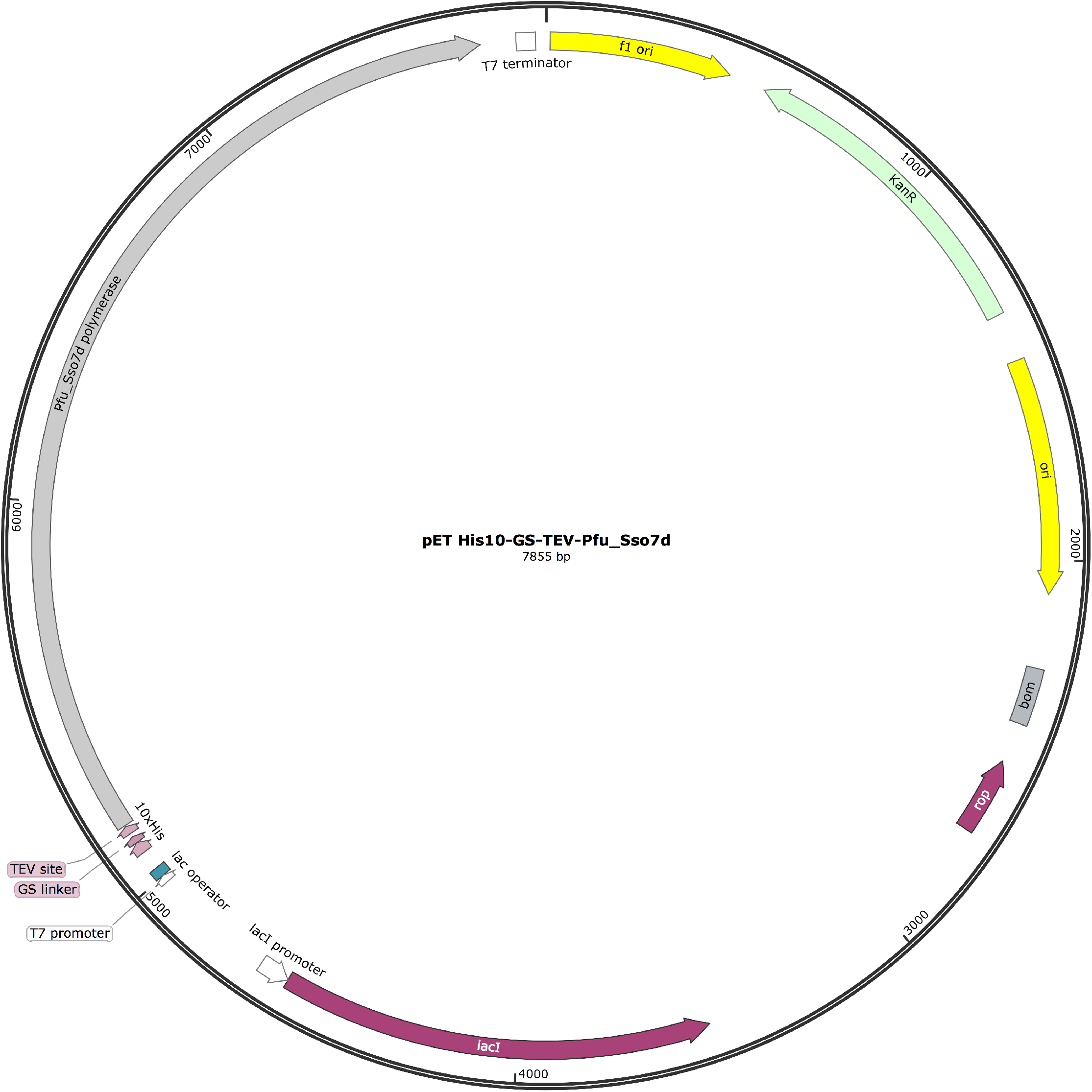
Plamid map of pET His10-GS-TEV-Pfu-Sso7d.

**Figure 3.**
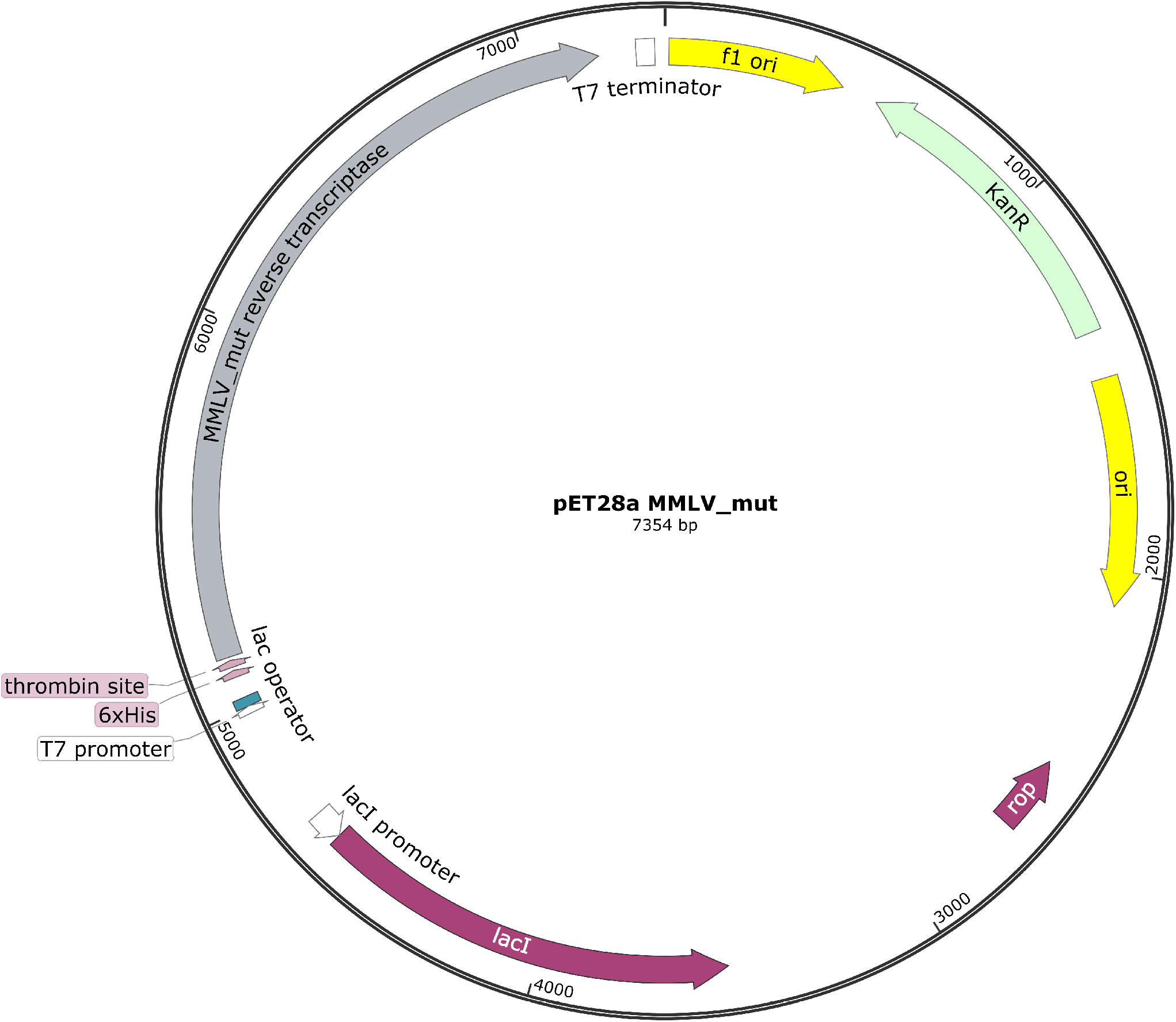
Plamid map of pET28a MMLV_mut.

### Supplementary File 3: Automation of the McQ reverse transcription step on the Biomek i7 liquid-handling platform

Reverse transcription (RT) is the major bottleneck in the McQ workflow. To increase throughput we implemented an automated RT protocol on the Beckman i7 automation system at the EMBL Genomics Core Facility. We believe that the method can easily be implemented on any liquid-handling system, using the original manual protocol as a guideline. Individual steps as programmed on the Biomek i7 are described in Figure 1, deck layout at start is shown in Figure 2, and chemistry setup in Figure 3. Table 1 lists required consumables for processing two plates. Master mixes are prepared in advance in 1.5 ml tubes and kept on ice until used. Lids have to be cut off tubes immediately prior to start of the protocol. The index master plate (containing the three target-specific barcoded primers and dNTPs in each well) is prepared in a PCR plate as for manual processing. All incubation steps requiring a thermal cycler are run off-deck to allow for parallel processing of several plates. The protocol starts with aliquoting the master-mixes from 1.5 ml tubes into chilled PCR plates. Reaction plates are set up using 10µl extracted RNA and 3µl index master mix, followed by off-deck incubation for RT primer annealing. After incubation plates are placed back on the deck and RT master mix (containing enzyme, buffer and additives) is added. Reverse transcription is performed off-deck, plates are placed back on deck for addition of Exonuclease I, followed by off-deck incubation for ExoI digest, after which the samples are ready for pooling and cleanup. The system prompts the user to clean the deck and shut down the instrument at the end of the protocol.

**Figure 1:**
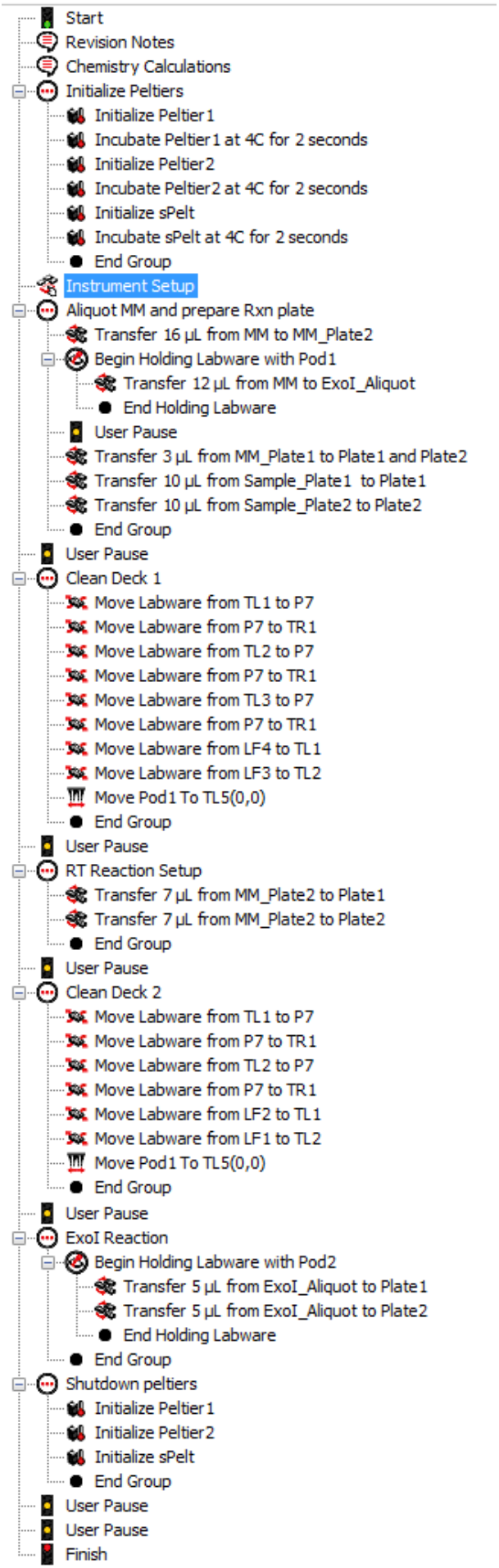
Workflow for automated reverse transcription implemented on the Biomek i7 platform. The figure shows a detailed step-by-step workflow of the protocol used for reverse transcription. Steps follow the manual protocol closely and use 10µl extracted RNA as input.

**Figure 2:**
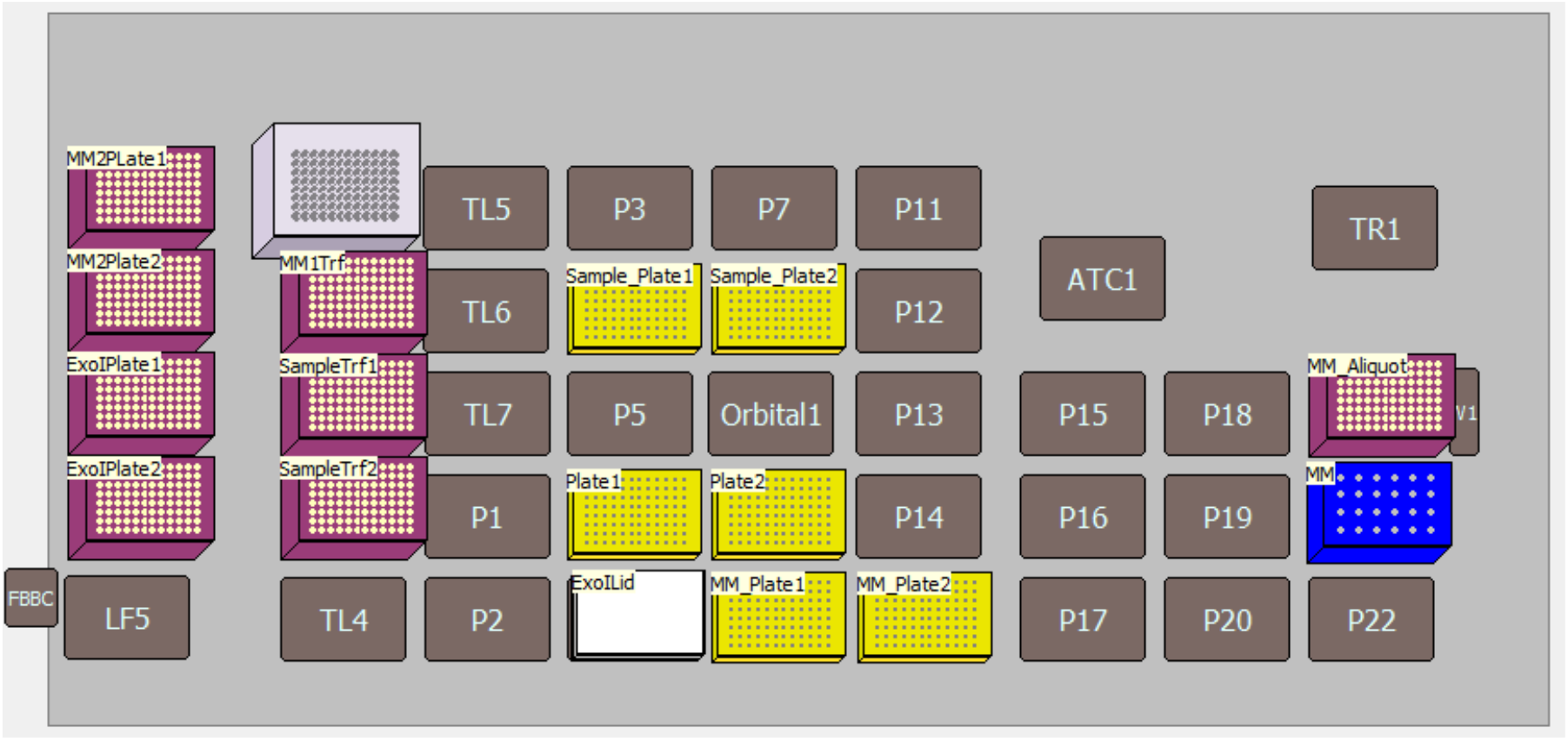
Deck-layout of the i7 system for reverse transcription. Pink boxes are 50ul Beckman tip boxes, yellow plates are BioRad Hard-Shell PCR Plates, the white lid is a single use plastic lid by Axon and the blue block is a chilled reagent block for 1.5ml DNA lo-bind Eppendorf tubes.

**Figure 3:**
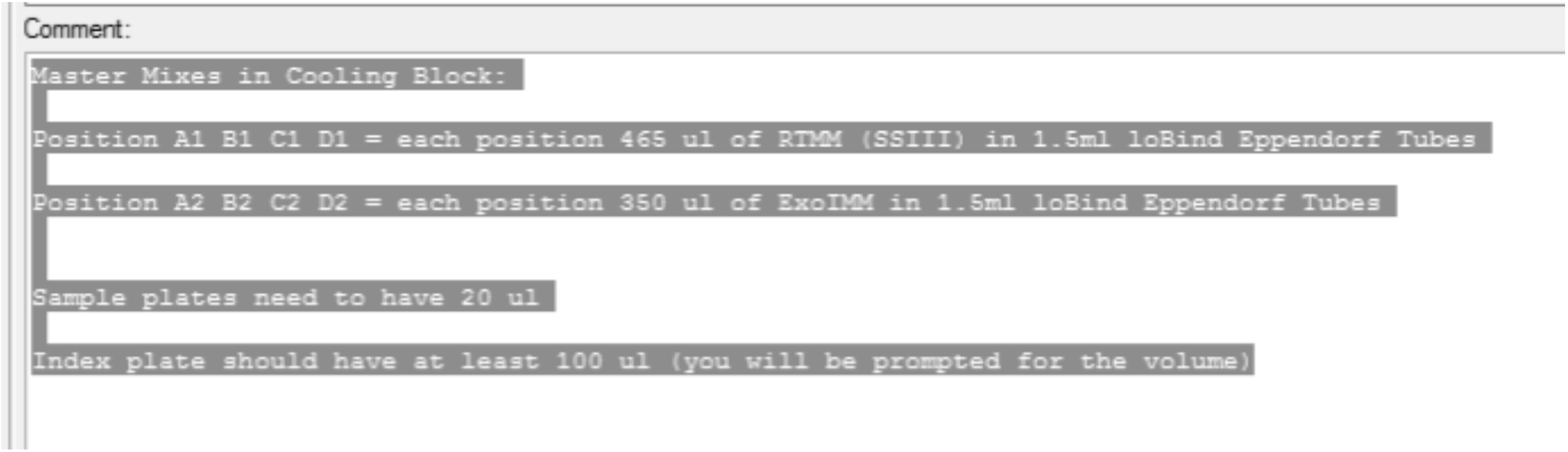
Chemistry Setup. Setup of the chemistry in the cooling-rack, as well as input criteria for sample and index plate.

**Table 1:** Consumables.

| *Item* | *Provider* | *Order #* | *Number used (per 2 plates)* |
| --- | --- | --- | --- |
| 50 ul tips | Beckman | B85888 | 8 |
| PCR plates | BioRad | HSP-9621 | 7 |
| Plastic lids | Axon | 4TI-0290 | 1 |
| Eppendorf tubes | Eppendorf | 30108051 | 12 |

### McQ step by step protocol for 96 samples

#### Critical notes before starting

- *Pipet everything on ice!*
- *Prepare the index master mix plate prior to starting, this plate can be re-used for processing several 96-well plates. We recommend making a large amount of stock once, in an* ***area/room free of potential virus*** *contamination (no synthetic virus material or swab samples handled, and using no equipment or reagents that have been used for processing such samples), and aliquoting the master mix according to anticipated number of plates that are processed per run. Discard the remainder of the aliquot to avoid carrying molecular contamination into following runs*.
- *Prepare RT Buffer V7, ExoI dilution buffer, and Pfu-Sso7d HF buffer V5*.
- *Trehalose in RT Buffer V7 will fall out of solution at low temperatures. Trehalose can be re-solubilized by incubation at 40°C in a water bath for 30 minutes (intermittent shaking)*.

#### Preparation step – Make an index master mix plate

Make an index master mix plate in a PCR plate. This plate can be used for processing several 96-well plates. If oligos are ordered at 100µM first make dilutions of the oligo plate to 10µM. See comments above for aliquoting index master mix.

Add per well:

| Reagent | 1X (µl) |
| --- | --- |
| dNTPs 10 mM | 1 |
| Nuclease-free water (NFW) | 1 |
| Indexed nsp14 RT primer 10 uM | 0.4 |
| Indexed E RT primer 10 uM | 0.4 |
| Indexed RP RT primer 10 uM | 0.2 |

#### Step 1a - Reverse Transcription (manual processing)

*Pipet everything on ice!*

*Prepare RT enzyme mix*.

Add 3 ul index master mix into 96-well plates

Add 10 ul sample

***Incubate 65°C 5 min***

***Place on ice 1 min***

*Note: Take the plate out of the cycler after 5 min and put it on ice immediately, do not let it cool down to 10°C in cycler*

RT enzyme mix:

| Reagent | 1X (µl) |
| --- | --- |
| 5X MMLV_mut buffer V7 | 4 |
| NFW | 0.5 |
| DTT 100 mM | 1 |
| RNAse inhibitor | 0.5 |
| MMLV_mut 0.1 mg/ml stock | 1 |

*Note: Prepare the master mix and aliquot it into 8-well PCR strips, calculating enough mix to process 1.3X the number of samples due to the high viscosity of the RT enzyme mix.*

Add 7 ul master mix to samples

***55°C 30 min***

***70°C 15 min***

Dilute Exonuclease I (ExoI) 1:1 in ExoI dilution buffer (to reduce viscosity), and add 2 µl diluted ExoI to each well (helps with viscosity)

***37°C 30 min***

***85°C 5 min***

#### Step 1b - Reverse Transcription (Biomek i7)

*Pipet everything on ice!*

Prepare index master mix in yellow Biorad plates (see above) Prepare samples in yellow Biorad plates (20µl volume)

Prepare 4 tubes of enzyme master mix (per tube 455 ul): 5X V7 buffer 260 ul

NFW 32.5 ul

DTT 100 mM 65 ul

RRI 32.5 ul

SSIII 0.1 mg/ml 65 ul

Prepare 4 tubes of ExoI master mix (per tube 350 ul): ExoI 70 ul

ExoI dilution buffer 280 ul

Run RT script.

#### Step 2 – pool and clean cDNA

- Combine 10 µl of each RT reaction into a single 1.5 ml tube (960µl total)
- Perform 1X (v/v) SPRI cleanup: Add 30 ul SPRI beads to pool, then add 930 ul SPRI buffer (without beads) for a total buffer to sample volume of 1X. Extra buffer can be prepared or obtained from taking the supernatant of 930µl of beads bound on a magnet.
- Mix well and incubate 5 min at room temperature
- Transfer tube to magnet and let bind for 5 minutes (until the solution is completely clear). This can be aided by pipetting up and down several times while the tube is bound on the magnet.
- Remove the buffer and do two washes with 80% ethanol (make sure beads are completely covered), completely remove residual ethanol, and take the tube off the magnet for drying (∼1 minute)
- Add 30 µl NFW and resuspend beads, transfer the sample to a PCR tube and incubate for 1-2 minutes
- Transfer to a PCR-tube sized magnet, let beads bind for 1-2 minutes, and transfer eluate to a fresh tube

#### Step 3 - PCR1

*Keep mastermix and samples on ice during pipetting as this helps reduce primer dimer. Keep reactions on ice and put directly into pre-warmed block (98°C) when using Pfu-Sso7d*.

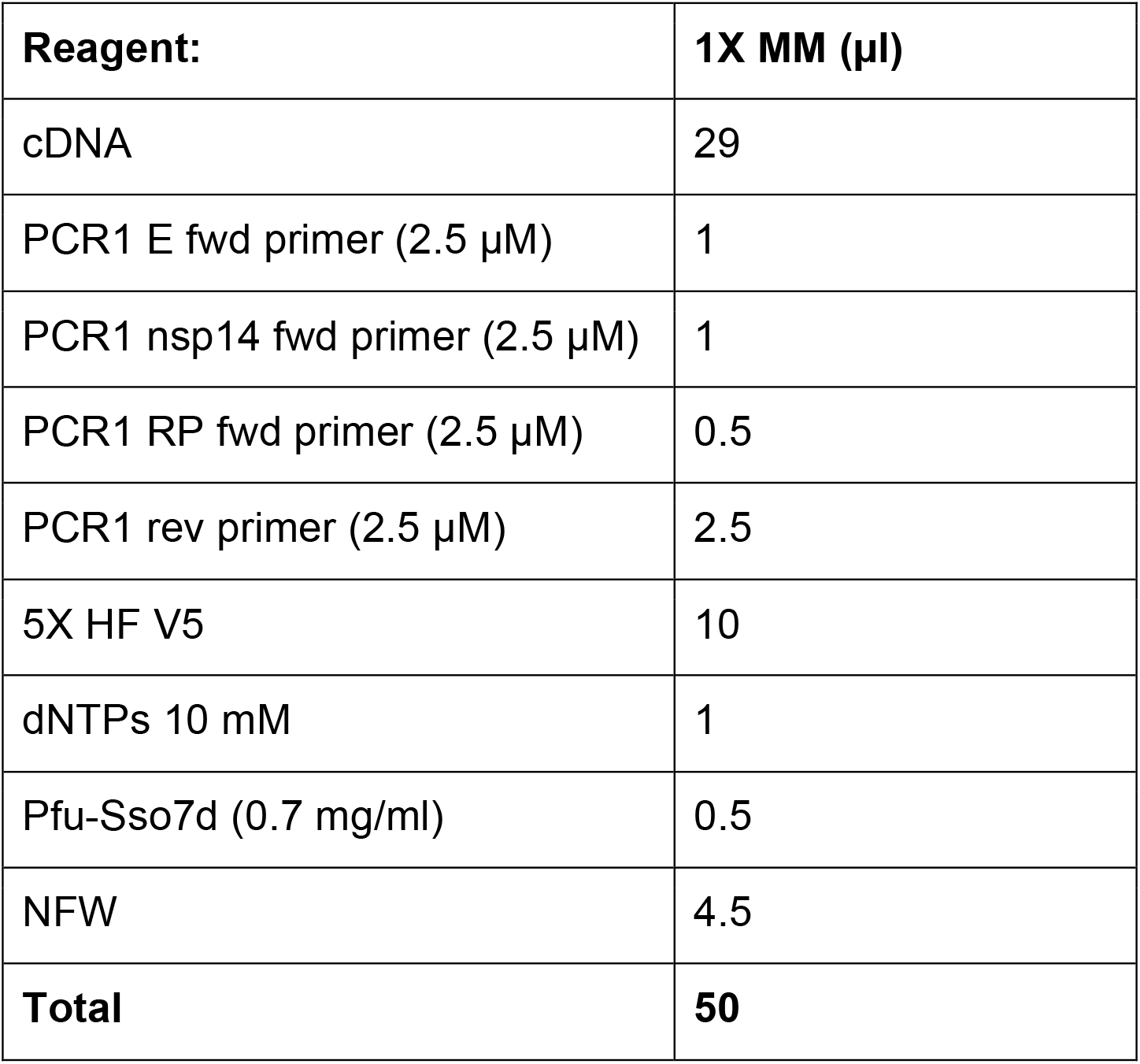

Add 21 ul master mix to cDNA

Cycler (∼55 min):

98°C 30 s

98°C 10 s

62°C 20 s

72°C 20 s

72°C 5 min 30X

#### Step 4 – SPRI cleanup PCR1

1.8X SPRI bead cleanup. Add 30 ul beads and 60 ul SPRI buffer to sample (total buffer volume 90 µl), and perform cleanup in PCR tubes, following protocol described in Step 2. Elute in 15 µl NFW.

#### Step 5 - PCR2

*Keep mastermix and samples on ice during pipetting as this helps reduce primer dimer. Keep reactions on ice and put directly into pre-warmed block (98°C)*.

Master mix:

| Reagent | 1X<br>(µl) |
| --- | --- |
| PCR1 | 3 |
| 5X HF V5 | 5 |
| dNTPs 10mM | 0.5 |
| Pfu-Sso7d (0.7 mg/ml) | 0.25 |
| NFW | 14.2<br>5 |

Add 1 µl 10 µM indexed P5

Add 1 µl 10 µM indexed P7

Cycler (∼25 min): 98°C 2 min

98°C 20 s

62°C 30 s

72°C 15 s

72°C 2 min

10X

#### Step 6 – ExoI digest to remove excess index primer (optional)

Add additional ExoI step to reduce contamination due to index hopping during sequencing caused by leftover indexed primers.

Add 1 µl ExoI directly into amplified PCR 2 and incubate 30 min at 37°C followed by 5 min at 85°C.

#### Step 7 – SPRI cleanup PCR2

1X SPRI bead cleanup. Use 25 µl beads and follow protocol in Step 3, elute in 15 µl NFW.

#### Step 8 – Pooling and QC

Dilute samples 1:10 and measure concentration with Qubit hsDNA assay. Pool equal amounts (ng) of each pool (adjust according to lowest concentration), check quality on Bioanalyzer, and submit for 75SE sequencing.

#### Buffers

| **1x MMLV_mut Reaction Buffer V7** | **5 x MMLV_mut Reaction Buffer V7** |
| --- | --- |
| 25 mM Tris-HCl pH 8.3 @ 25°C<br>25 mM MOPS pH 7.9 @ 25°C<br>60 mM KCl<br>4 mM MgCl<sub>2</sub><br>200 mM Trehalose<br>5% Glycerol<br>0.006% Igepal CA-630 | 125 mM Tris-HCl pH 8.3 @ 25°C<br>125 mM MOPS pH 7.9 @ 25°C<br>300 mM KCl<br>20 mM MgCl<sub>2</sub><br>1000 mM Trehalose<br>25% Glycerol<br>0.03% Igepal CA-630 |

*Note: Trehalose is on the limit of solubilization in the 5X buffer due to the addition of 25% Glycerol and might require heating at 40°C to promote complete solubilization.*
*Buffer V7 is adapted from (https://pipettejockey.com/2019/07/15/mashup-rt-update-2-our-take-on-purification-activity-assays-buffer-optimization/) by addition of Trehalose.*

| **1x Pfu-Sso7d HF buffer V5** | **5x Pfu-Sso7d HF buffer V5** |
| --- | --- |
| 10 mM POPSO (pH 8.8 @ 25°C) | 50 mM POPSO (pH 8.8 @ 25°C) |
| 80 mM KCl | 400 mM KCl |
| 16 mM (NH<sub>4</sub>)<sub>2</sub>SO<sub>4</sub> | 80 mM (NH<sub>4</sub>)<sub>2</sub>SO<sub>4</sub> |
| 1.5 mM MgCl<sub>2</sub> | 7.5 mM MgCl<sub>2</sub> |
| 10 mM Arginine | 50 mM Arginine |
| 200 mM Trehalose | 1000 mM Trehalose |
| 100 mM Sarcosine | 500 mM Sarcosine |
| 0.1% Tween-20 | 0.5% Tween-20 |

*Note: HF buffer V5 is from Alexander Klenov*
[\(https://pipettejockey.com/2017/08/18/purifying-commonly-used-enzymes-homebrew-phusion/\)](https://pipettejockey.com/2017/08/18/purifying-commonly-used-enzymes-homebrew-phusion/)

### Exo I dilution buffer

10 mM Tris-HCl pH 7.4 @ 25°C

0.1 mM EDTA 1 mM DTT

0.25 M NaCl 10% Glycerol 200 µg/ml BSA

## Notes

### Competing Interest Statement

The authors have declared no competing interest.

### Funding Statement

D.B., M.F. and J. K. were supported by the EMBL International PhD program. D.B. was supported by a Darwin Trust fellowship. Research in the laboratory of A.R.K and L.S. was supported by core funding of the European Molecular Biology Laboratory. M.K. and H.G.K received funding through a research grant from the state of Baden-Wuerttemberg (MWK).

### Author Declarations

EMBL Bioethics Internal Advisory Committee (BIAC) & Ethics Committee of the Medical Faculty of Heidelberg

